# The differential effects of common and rare genetic variants on cognitive performance across development

**DOI:** 10.1101/2024.09.04.24313061

**Authors:** Daniel S. Malawsky, Mahmoud Koko, Petr Danacek, Wei Huang, Olivia Wootton, Qin Qin Huang, Emma E. Wade, Sarah J. Lindsay, Rosalind Arden, Matthew E. Hurles, Hilary C. Martin

## Abstract

Common and rare genetic variants that impact adult cognitive performance also predispose to rare neurodevelopmental conditions involving cognitive deficits in children. However, their influence on cognition across early life remains poorly understood. Here, we investigate the contribution of common genome-wide and rare exonic variation to cognitive performance across childhood and adolescence primarily using the Avon Longitudinal Study of Parents and Children (n=6,495 unrelated children). We show that the effect of common variants associated with educational attainment and adult cognitive performance increases as children age. Conversely, the negative effect of deleterious rare variants attenuates with age. Using trio analyses, we show that these age-related trends are driven by direct genetic effects on the individual who carries these variants rather than indirect genetic effects mediated via the family environment. We further find that the increasing effects of common variants are stronger in individuals at the upper end of the phenotype distribution, whereas the attenuating effects of rare variants are stronger in those at the lower end. Concordant results were observed in the Millenium Cohort Study (5,920 children) and UK Biobank (101,232 adults). The effects of common and rare genetic variation on childhood cognitive performance are broadly comparable in magnitude to those of other factors such as parental educational attainment, maternal illness and preterm birth. The effects of maternal illness and preterm birth on childhood cognitive performance also attenuate with age, whereas the effect of parental educational attainment does not. We show that the relative contribution of these various factors differs depending on whether one considers their contribution to phenotypic variance across the entire population or to the risk of poor outcomes. Our findings may help explain the apparent incomplete penetrance of rare damaging variants associated with neurodevelopmental conditions. More generally, they also show the importance of studying dynamic genetic influences across the life course and their differential effects across the phenotype distribution.

## Main text

Cognitive ability is an important predictor of life outcomes such as health, education, occupation, and mortality^1–6^. It is influenced both by genetic and environmental factors^7^, including extrinsic factors that have both genetic and environmental components such as parental educational attainment (EA), gestational age at birth, and maternal illness during pregnancy^8–10^. Results from twin studies show that the heritability of cognitive ability increases throughout development, from ∼0.2 in infancy to ∼0.6 in adulthood^11^. In large meta-analysis of adults, the heritability of cognitive performance attributable to common single-nucleotide polymorphisms (the ‘SNP heritability’) was estimated to be 0.20 ^12^. The SNP heritability is likely to be explained both by direct genetic effects^12,13^ (the effects of genetic variants in an individual on that individual’s own phenotype) and by a variety of other factors. These other factors include the indirect effects of “genetic nurture” (a phenomenon whereby an individual’s rearing environment is influenced by their relatives’ genetic make-up), parental assortment (i.e. phenotypic correlation between partners, commonly known as ‘assortative mating’), and uncontrolled population stratification in genome-wide association studies (GWASs)^14–17^. The relative contribution of genetic nurture, parental assortment and population stratification to genetic associations is still a matter of debate, and likely differs between cognitive performance and related traits such as EA and academic achievement^14,18–21^. However, the role of direct versus indirect genetic effects on cognitive and academic performance across different timepoints in development is under-studied, and we explore this in this work.

It has become increasingly clear that the genetics of rare neurodevelopmental conditions (NDCs) involving cognitive impairment overlap with genetic factors impacting cognitive ability in the general population. While Mendelian-acting rare coding variants play a large role in NDCs, explaining ∼50% of probands in the Deciphering Developmental Disorders (DDD) study^22–24^, common variants explain ∼10% of variation in risk of these conditions on the liability scale^25,26^. This common variant risk is negatively genetically correlated with EA and adult cognitive performance in the general population^25,26^. Additionally, rare damaging coding variants in NDC-associated genes are associated with lower fluid intelligence and lower EA in UK Biobank^27–29^. Estimates suggest rare coding variants exome-wide explain 1% of the variance in adult cognitive performance^30^.

Multiple lines of evidence from both clinical^31–33^ and population cohorts^29,34^ suggest that some rare variants conferring risk of NDCs are often shared between affected children and their seemingly unaffected parents. This so-called ‘incomplete penetrance’ is seen both for large copy-number variants^32^ and for predicted loss of function variants (pLoFs) and damaging missense variants in genes in which such variants have been under negative selection throughout recent human evolution (‘constrained genes’^35^)^33^. It is frequently assumed that multiple genetic and environmental factors contribute to an underlying liability for NDCs, and children are diagnosed if they cross some phenotypic threshold^36^. Under this model, incomplete penetrance of rare variants is expected if they increase the mean of the liability distribution in carriers to a point at which some but not all cross the threshold for diagnosis. It could also occur if the variants increase the *variance* in liability even without a change in mean liability of carriers, by pushing more individuals to the distribution extremes. Polygenic background affects liability^25,26^ and has been hypothesized to modify the penetrance of these rare variants; there is evidence of this from the general population^29^, but recent within-family analyses in NDC cohorts failed to find support for it^26^. Another cause of apparent incomplete penetrance could be time-varying effects i.e., rare variants (or at least a subset of them) influences cognitive ability in childhood more than they do in adulthood. We investigate this here.

In this work, we sought to dissect the contribution of genome-wide common variants and rare exonic variants to cognitive performance across childhood and adolescence, using genotype data and new exome-sequence data (N=16,103 children and 10,140 parents) from two British birth cohorts, the Avon Longitudinal Study of Parents and Children (ALSPAC)^37^ and the Millenium Cohort Study (MCS)^38^. Our first aim was to examine the effects of genetic measures on average cognitive performance and academic achievement as children age, then test whether these change across time and the extent to which they are due to direct genetic effects. Our second aim was to explore whether these genetic measures have differential effects at the tails of the phenotype distribution (in other words, whether they impact the variance in cognitive ability). Finally, our third aim was to compare these genetic effects to those of parental educational attainment and perinatal exposures, and to explore whether they are robust to controlling for these other measures. Our results illuminate the dynamic genetic architecture of cognitive performance across development and shed light on the factors that best predict cognitive impairment in childhood as opposed to average cognitive performance in adulthood.

## Results

In this work, we used measurements of cognitive performance and/or school achievement from three cohorts: ALSPAC, MCS and UK Biobank. Our primary analyses were based on measures of IQ from ALSPAC that were collected at ages 4 (n=1,012), 8 (n=7,347), and 16 (n=5,270). These IQ tests have good psychometric properties^39^ and are longitudinally invariant, meaning that they measure the same latent construct across ages^40^, making them suitable for longitudinal analyses. Since phenotypic missingness can induce ascertainment biases and reduce power, we imputed missing IQ values across ages by leveraging the lower missingness of other cognitive and behavioral tests and demographic variables (Supplementary Note 1; Extended Data Figure 1). We carried out analyses on both the observed IQ values (“pre-imputation”) and on the observed and imputed IQ values combined (“post-imputation”). Results were largely concordant between these unless specifically noted otherwise.

### Influence of common variants on cognitive performance across development

To assess the overall contribution of common variants to cognitive development, we first considered the heritabilities and genetic correlations of IQ measured at different ages in ALSPAC, using 6,495 unrelated children with SNP genotype data, genetically-inferred European ancestry and observed or imputed IQ values. Using GREML-LDMS^41^, we estimated that the heritability of IQ increased between ages 4 to 16 from 0.46 to 0.56 (post-imputation), though the increase was not significant (p=0.053) (Table S2). The pairwise genetic correlations between IQ measures from different ages were all indistinguishable from 1 (Table S2), suggesting the common variant genetic architecture is largely stable across development. These results are consistent with previous studies^42,43^.

We then considered the associations between IQ and polygenic indices (PGIs), which capture the effect of common SNPs ascertained for their association with a given trait. We first considered a PGI for EA (PGI_EA_). EA is known to have a cognitive component as well as a non-cognitive component which may capture factors such as personality traits and socioeconomic status that affect people’s ability to progress through education^44^. Genetic effects on these two components have been previously derived from GWASs in adults using Genomic Structural Equation Modeling^44^, so we also included PGIs representing each of them (PGI_Cog_ and PGI_NonCog_). For each PGI, we modeled IQ across all ages as the outcome of individual-specific random effects and several fixed effects, namely age (relative to age 4), the PGI, and their interaction, with sex and genetic principal components as covariates. With this mixed-effects linear model, we were able to estimate two parameters of interest: the effect of the PGI at baseline i.e. at age 4 (the “main effect”, estimated leveraging the data at all ages) and whether the effect changed with age (the “interaction effect”; see Box 1). We detected significant (p<.05/3) positive main effects for all PGIs, as expected (“population model” in Figure 1A, Table S4). Importantly, we found that common variants ascertained for their association with cognitive performance and final educational attainment in adults explain more variance in IQ as children age: there were significant positive PGI-by-age interaction effects for all PGIs post-imputation (Figure 1A) as well as for PGI_EA_ pre-imputation (Table S4).

**Figure 1.**
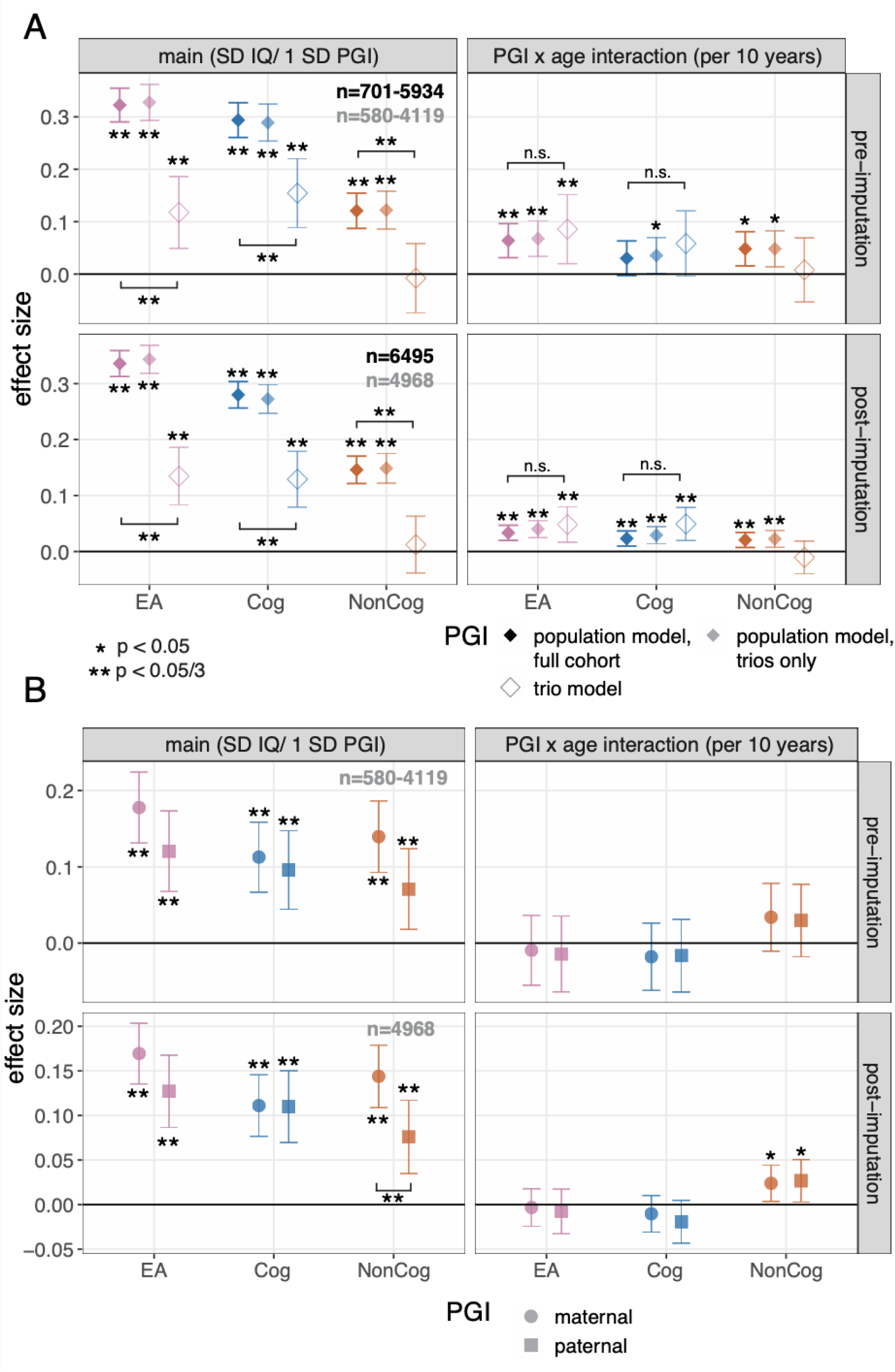
Association between PGIs and IQ across ages. Standardized effects and 95% confidence intervals estimated for the main effects and PGI-by-age-interaction effects for each PGI, either pre-(top) and post-imputation (bottom) A) Results for the children’s PGIs not controlling (i.e. ‘population model’) and controlling (right) for parental PGIs (i.e. trio model). Population effect sizes are shown estimated in the full sample (opaque) and in the subset of children with parental PGIs (translucent), with the corresponding sample sizes shown in black and gray text respectively. B) Results for the parental PGIs from the trio model. SD: standard deviation; PGI: polygenic index; EA: educational attainment; Cog: cognitive component of EA from ^19^; NonCog: non-cognitive component of EA from ^19^. The square brackets indicate significant comparisons highlighted in the text (z tests).

These associations between children’s PGIs and IQ, which we term ‘population effects’, likely reflect a combination of direct genetic effects and other correlates of the child’s genotype including the influence of parental genotypes on the child’s IQ (“genetic nurture”) and potential confounding due to population stratification and parental assortment. We next sought to specifically estimate the direct genetic effects of PGIs on IQ across development. We repeated the above analyses adding parental PGIs as covariates in a subset of 4,968 unrelated parent-offspring trios for which we measured parental genotypes or could infer them via Mendelian imputation^20^ (“trio model”; see Methods). In this model, the coefficient on the child’s PGI provides an estimate of the direct genetic effects^14^, while the coefficient on the parents’ PGIs represents the association between the child’s phenotype and alleles in the parents that are not transmitted to the child. Our results for PGI_NonCog_ suggested that its population effect reflects genetic nurture or potential confounding, since its direct effect was not significantly different from 0 (Figure 1A). In contrast, PGI_EA_ and PGI_Cog_ showed significant direct main effects on IQ, though weaker than the population effects (p<0.005, z test for difference between direct and population main effects) (Figure 1A). The increasing effects of PGI_EA_ and PGI_Cog w_ith age that we detected in the population model appear to reflect direct genetic effects, since the PGI-by-age interaction effects were significant and positive in the trio model (p<.05/3) and statistically indistinguishable from the population effects (p>0.05, z test). The cognitive component of EA fully captured the direct genetic effect of PGI_EA_ on IQ: we found no significant differences between the direct effects for PGI_EA_ and PGI_Cog_ (p>0.05 for difference in the main and interaction effects, z test).

In the trio model, all parental PGIs had significant main effects, indicating that genetic nurture and/or confounding effects captured by the parental PGIs are associated with childhood IQ (Figure 1B, Table S4). Only the parental alleles in PGI_NonCog_ showed evidence for a change with age. The effects captured by parental PGI_NonCog_ were stronger in mothers than fathers (difference in main effects post-imputation = 0.067, p=0.014, z test), but both effects increased with age to a similar degree (Figure 1B bottom right panel). These associations between children’s IQ and parental non-transmitted alleles associated with the non-cognitive component of EA could be because parenting behaviors correlated with these alleles increase children’s cognitive ability, or due to cross-trait parental assortment based on the cognitive and non-cognitive components of EA. The significant difference between the maternal and paternal effects may suggest that there is at least some ‘genetic nurture’ component which differs between parents.

#### Box 1

##### Genetic effects estimated in this study

We estimate a variety of genetic effects from mixed-effects models (see Methods section on “Associations between genetic measures and traits”), including either main or interaction effects combined with either population effects, direct effects or effects of non-transmitted parental alleles.

###### Population main effect

The effect of a child’s genetic score on the phenotype at the first measurement time point (e.g., age 4 in the ALSPAC IQ analysis), but leveraging information from all time points to estimate this (see Methods). This effect is estimated without adjusting for parental genetic scores in the regression, and reflects a combination of the effects of the child’s genetic score on their own phenotype, genetic nurture, parental assortment, and uncontrolled population stratification. It does **not** have a causal interpretation.

###### Population interaction effect

The *change* in the population effect of a genetic score on the phenotype with age. e.g., If the main effect is positive and the interaction effect is positive, this indicates that the effect of the genetic score increases with age, whereas if the main effect is negative and the interaction effect is positive, this indicates that the effect attenuates with age.

###### Direct main effect

The effect of a child’s genetic score on the phenotype at the first measurement time point, adjusting for the parental genetic scores (i.e., a trio model). This effect isolates the influence of the child’s own genetic score on their own phenotype and has a causal interpretation (see section IV in ^45^ for further discussion).

###### Direct interaction effect

The *change* in the direct effect of a genetic score on the phenotype with age. The interpretation otherwise mirrors that of the population interaction effect.

###### Effects of non-transmitted alleles

These are the effects of the parental genetic scores on the phenotype, estimated in the trio model; in this model, the direct effect of transmitted alleles is captured by the coefficient on the child’s score, whereas the coefficients on the parents’ scores are mathematically equivalent to the effect of the non-transmitted alleles in the parents. These effects reflect a combination of genetic nurture, parental assortment, and uncontrolled population stratification. They do **not** have a causal interpretation.

Note that all of these effects are estimated both on the mean of the phenotype with linear regression (Figures 1, 2, 4A), and also on quantiles of the phenotype (e.g., the 5th percentile) with quantile regression (Figure 3, 4B).

**Figure 2.**
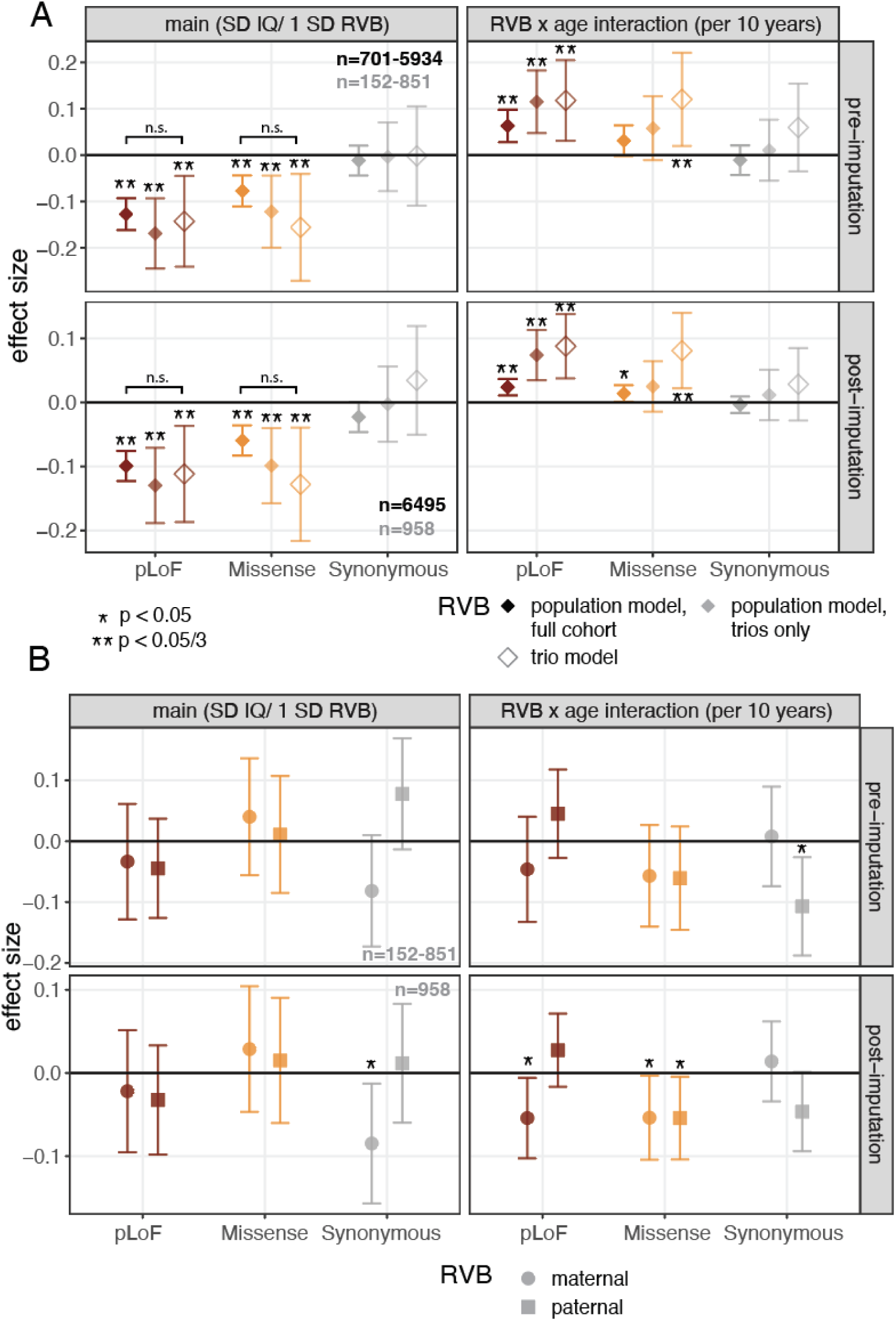
Association between rare variant burden (RVB) and IQ across ages. Standardized effects and 95% confidence intervals estimated for the main effects and RVB-by-age-interaction effects for RVB calculated with three different consequence classes (pLoF, missense or synonymous), either pre-(top) and post-imputation (bottom). A) Results for the children’s RVBs not controlling (i.e. ‘population model’) and controlling (right) for parental RVBs (i.e. trio model). Population effect sizes are shown estimated in the full sample (opaque) and in the subset of children with parental RVBs (translucent), with the corresponding sample sizes shown in black and gray text respectively. B) Results for the parental RVBs from the trio model. The square brackets indicate comparisons highlighted in the text (z tests).

**Figure 3.**
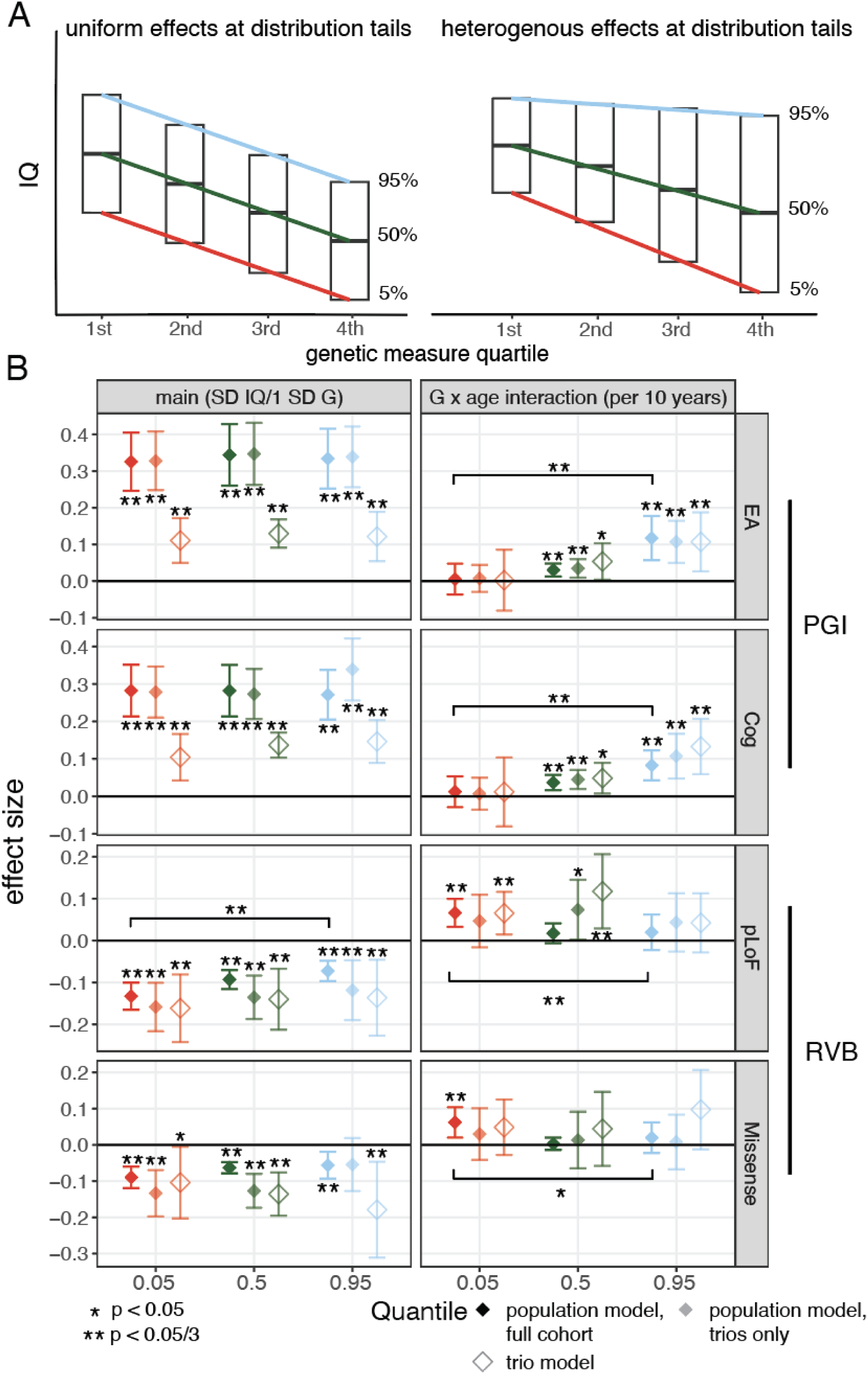
Influence of common and rare variants on the tails of the IQ distribution, using IQ measures post-imputation in ALSPAC. A) Schematic showing two scenarios where a genetic measure is associated with IQ and has uniform effects across the IQ distribution (left) versus a scenario in which the genetic measure has heterogeneous effects on IQ across the IQ distribution (right). B) Standardized effects and 95% confidence intervals for quantile regression of the 5th, 50th, and 95th percentile estimated from mixed-effects modeling with post-imputation IQ at ages 4, 8, and 16 for EA and EA-cog PGIs before (left) and after (right) controlling for parental genetic measures. C) Same as (B) using RVB_pLoF_ and RVB as genetic measures. See Figure S3 for results from this mixed-effects model pre-imputation, and Figure S2 and Figure S4 for cross-sectional estimates at each age. The square brackets indicate significant comparisons highlighted in the text (z tests).

**Figure 4.**
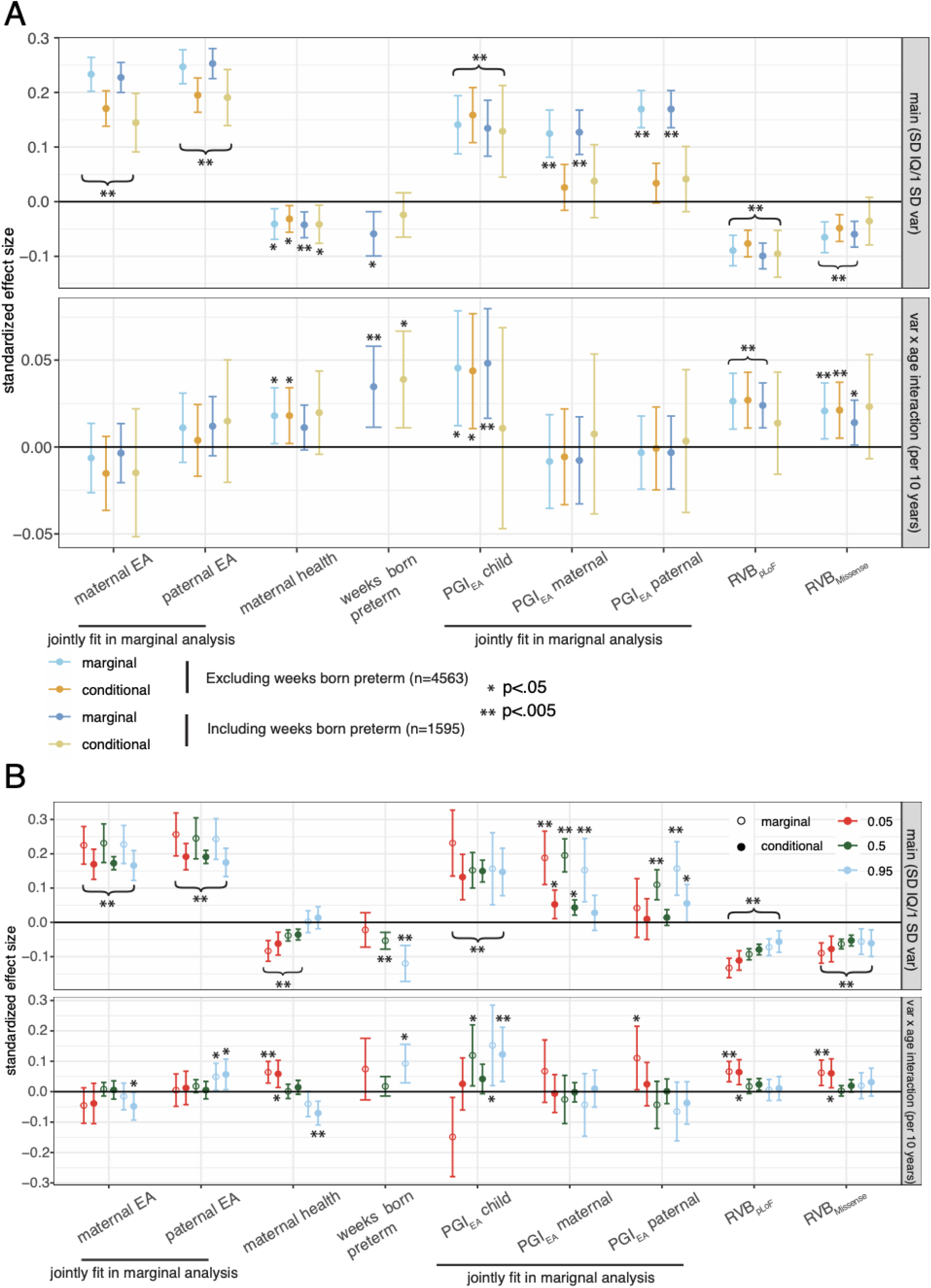
Associations between genetic and other factors and IQ across ages in ALSPAC. A) Standardized effects and 95% confidence intervals for the main effects (top) and measure-by-age-interaction effects (bottom) obtained from mixed-effects models fitted for IQ post-imputation. These were obtained from four models, as indicated in the key: marginal associations from models in which only the indicated variable(s) were included in the model (in addition to standard covariates), and conditional associations from models in which all the variables were included in the same model. Note that we estimated the marginal and conditional effects either using only individuals who had gestational age information (n=1,595) or all individuals (n=4,563), dropping the “weeks born preterm” variable in the latter case. See Methods for details. B) Standardized effects and 95% confidence intervals for the main effects (top) and measure-by-age-interaction effects (bottom) for quantile regression of the 5th, 50th, and 95th percentile estimated from mixed-effects quantile regression modeling for IQ post-imputation using both marginal and conditional associations as in (A). In (B), marginal effect sizes for the child’s PGI_EA,_ RVB_pLoF,_ and RVB_Missense,_ which were previously shown in Figures 1 and 2 are replotted to facilitate comparisons of effect size with the conditional analysis results. Asterisks indicate whether the estimate is significantly different from 0; note that curly brackets with asterisks indicate that the estimates spanned by the bracket are all significant.

Repeating these PGI analyses using a measure of academic performance in ALSPAC gave similar results (Supplementary Note 2; Extended Data Figure 2, Table S5).

### Influence of rare variant burden on cognitive performance across development

In order to query the effects of rare protein-altering variants, we generated new exome sequencing data on ALSPAC (8,436 children and 3,215 parents) and MCS (7,667 children and 6,925 parents) to supplement the SNP genotype and longitudinal phenotype data already available. We first used these to assess the associations between IQ and rare variant burden (RVB) in ALSPAC. We quantified RVB as the sum of gene-specific selection coefficients^46^ (reflecting evolutionary constraint, or negative selection) for genes in which an individual carries a rare predicted loss-of-function or deleterious missense variant (RVB_pLoF_, RVB_Missense_; within-sample minor allele frequency, MAF <0.1%; see Methods), and, as a negative control, calculated an equivalent score for synonymous variants (RVB_Synonymous_).

Deleterious rare variant burden measured by RVB_pLoF_ and RVB_Missense_ was significantly negatively associated with IQ (main effects p<.05/3), whereas the burden of rare synonymous variants was not (Figure 2A left, Table S6). As expected, the unstandardized effect of deleterious LoF variants was stronger than that of deleterious missense variants, with the main effect of RVB_pLoF_ being 3.2-times larger than that of RVB_Missense_ (p=4×10^-7^, z test) (Extended Data Figure 3). In contrast to the results seen above for common variants, the effect of deleterious rare variants attenuated with age (p<.05/3 for RVB_pLoF_, and p=0.033 for RVB_Missense_ post-imputation) (Figure 2A right). Repeating these analyses using an academic performance measure in ALSPAC yielded similar results (Supplementary Note 2).

Next we considered the effects of RVB_pLoF_ calculated for specific gene sets. We found that genes prioritized via common variant-based GWAS of educational attainment^47^ showed a stronger association with IQ in childhood than expected for similarly evolutionarily constrained genes, highlighting the convergence of common and rare variant associations for these related phenotypes on the same genes (Supplementary Note 3, Extended Data Figure 5). We also found suggestive evidence that the attenuation of the effect of RVB_pLoF_ on IQ with age was strongest in genes that showed preferential expression in the prenatal rather than postnatal brain ^48^ (Supplementary Note 3, Extended Data Figure 5A), although this was less clear when examining academic performance (Figure S5).

We then sought to estimate the direct genetic effects of deleterious rare variants on IQ across development in a subset of 958 unrelated exome-sequenced trios by adding parental RVBs as covariates. Our findings suggested that the population effects are dominated by the direct effects of rare variants, in contrast to what we observed for PGIs: the estimated population and direct main effects were not significantly different from each other for either RVB_pLoF_ nor RVB_Missense_ (p>0.05, z test) (Figure 2A, Table S6) and no parental RVB measure was significantly associated with IQ after accounting for multiple testing (Figure 2B, Table S6), though this may be in part due to the smaller sample size. We also found that the direct effects of these rare variants significantly attenuated with age (p<.05/3 for RVB_pLoF_-by-age and RVB_Missense_-by-age interactions in the trio model). In summary, our results show that higher exome-wide burden of rare damaging variants is associated with lower IQ in childhood, though with attenuated effects at later age, and that this pattern is recapitulated when examining the direct genetic effects. When partitioning the effect of the burden of rare pLoFs in constrained genes into inherited variants versus *de novo* mutations, we estimated that the former explained 3.5-4 times more variance in IQ than the latter in ALSPAC (Supplementary Note 4; Extended Data Figure 6), with similar results when considering a composite measure of childhood cognitive performance in MCS (see Methods; Figure S1).

### Influence of genetic factors on the tails of the IQ distribution

Our results thus far consider genetic effects on mean IQ. However, we have yet to consider whether they differentially impact IQ at different quantiles in the IQ distribution. Importantly, deleterious rare variants in constrained genes are known to affect risk of neurodevelopmental conditions^22^, hinting that they may possibly have larger effects at the lower tail end of the IQ distribution. This would reflect a statistical phenomenon known as “heteroscedasticity”, in which, in a linear regression of phenotype on genotype, the variance of the residuals varies by genotype. To test whether there are heterogenous genetic effects at the tails of the IQ distribution, we used quantile regression to estimate the influence of PGIs and RVB on both the median IQ as well as the bottom and top 5th percentiles (Figure 3A), which roughly correspond to IQs of 75 and 125, respectively^39^.

First, we considered the influence of PGI_EA_ and PGI_Cog_, which had significant direct genetic effects on IQ in Figure 1. Broadly, our results suggested that the observed increase of these PGI effects on IQ with age was driven by increasing genetic effects in the upper half of the IQ distribution. Although the effects of the PGIs on these different quantiles in the IQ distribution were largely concordant at any given time point (Figure S2, Table S8), we found significant positive age interaction effects for both PGIs at the median and 95th quantile for population effects (p<.05/3), which were at least nominally significant when considering the direct effects (Figure 3B). Notably, there was no evidence for significant age interactions at the 5th percentile for either PGI. Repeating this analysis using academic performance in ALSPAC replicated the results, showing that the effect of PGI_EA o_nly significantly increased at the median and nominally at the 95th percentile (Extended Data Figure 7A). We observed concordant results when using cognitive ability measures from MCS and UK Biobank (Supplementary Note 5; Extended Data Figure 7B). In ALSPAC, parental PGI associations with IQ were largely uniform across the phenotype distribution and consistent across development (Extended Data Figure 8).

Secondly, we considered the influence of rare variant burden on these different quantiles of the IQ distribution at different ages in ALSPAC. Broadly, we found that the effects of deleterious rare variants were strongest on those with low IQ, and that these effect attenuate more markedly with age than those on the upper tail of the distribution, reducing the heterogeneity in effects across the phenotype distribution with time (Figure S4; Table S9). Notably, at age 4, the population effect of RVB_pLoF_ at the 5th percentile was 3.9-times greater than that at the 95th percentile (p=9×10^-5^, z test) and 2.2-times greater than at the median (p=7×10^-4^), whereas by age 16, the estimates at the different quantiles were not significantly different from one another. Put another way, this implies that, at age 4, the variance in IQ in people with high RVB_pLoF_ is greater than in those with lower RVB_pLoF_, but this is driven by there being more people with lower rather than higher IQ, as illustrated in the right-hand plot in Figure 3A; however, by age 16, RVB_pLoF_ shows uniform effects at the distribution tails. As a consequence of this, the significant attenuation of population effects with age was observed only at the 5th percentile and median, but not at the 95th percentile, for both RVB_pLoF_ and RVB_Missense_ (Figure 3B). This finding was recapitulated when examining direct genetic effects of RVB_pLoF_ although not RVB_Missense_ (Figure 3B). Although we had detected no significant effects of parental RVB on mean IQ in the trio model (Figure 2B), we detected a nominally significant negative effect of maternal RVB_pLoF_ on the bottom 5th percentile, suggesting that the effect of genetically-influenced parental behaviors on IQ may vary across the IQ distribution (Extended Data Figure 8). This is in line with our recent finding of a significant effect of non-transmitted rare damaging maternal alleles on risk of neurodevelopmental conditions (Extended Data Figure 10B in ^26^). Repeating the RVB_pLoF_ analysis on school grades in ALSPAC, we similarly find stronger associations between RVB_pLoF_ and academic performance at the 5th percentile that attenuate as children age (Extended Data Figure 7A). An analysis using cognitive ability measures from MCS and UK Biobank yielded concordant observations (Supplementary Note 5; Extended Data Figure 7B).

In summary, we show that the negative effects of RVB are strongest on the lower tail of the distribution of cognitive ability, and that these effects attenuate with age. These results stand in contrast to those for the PGIs for which the significant age interactions were seen only in the top half of the IQ distribution.

### Relative contribution of genetic and other exposures to IQ

Finally, we compared the longitudinal effects of common and rare genetic variants on IQ (Figures 1 and 2) to the effects of other factors that have previously been associated with children’s academic and cognitive outcomes (see Methods). We tested the effects of two perinatal factors, namely maternal illness during pregnancy and premature birth (as measured by the number of weeks born preterm), and the effect of realized parental educational attainment on IQ in ALSPAC. When fitting the variables separately (i.e., marginal associations shown in blue points in Figure 4A), we found strong positive effects of paternal and maternal EA (Figure 4A top) that were constant with age (Figure 4A bottom). In contrast, the perinatal factors were negatively associated with IQ (Figure 4A top) but their effects attenuated with age (Figure 4A bottom), consistent with previous findings using school grades in ALSPAC and MCS^49,50^. The standardized effects for these perinatal exposures were weaker than those for rare variants, with maternal illness and weeks born preterm explaining 0.18% and 0.35% of the variance in IQ at age 4 respectively, versus 1.40% for RVB_pLoF_ and RVB_Missense_ collectively (p=5.2×10^-4^ and 0.027, respectively) (Figure 4A top). In a joint model including parental EA, perinatal exposures and genetic scores (offspring and parental PGI_EA_ and the child’s RVB_pLoF_ and RVB_Missense_), most effect sizes changed minimally, with the exception of the parental PGI_EA_ effects which became non-significant. Importantly, the effects of the child’s genetic scores did not significantly change (Figure 4A top, Supplementary Note 6), and the direct effect of the child’s PGI_EA_ was similar to the effects of parental EA. Conclusions were similar when considering a composite measure of childhood cognitive performance in MCS (Extended Data Figure 10A). Collectively, these results show that the magnitude of effects of these genetic measures on childhood IQ is comparable to those of other well-established influences and that the changing effects with age persist after controlling for other environmental factors (Figure 4A bottom).

Given our observation above of the differential effects of genetic measures on the tails of the IQ distribution, we further considered the influence of parental EA and these perinatal exposures on the different quantiles of the distribution, fitting models for each additional variable independently as well as joint models as mentioned above (Figure 4B; Extended Data Figure 10B; Supplementary Note 6). Our conclusions from the marginal analysis in Figure 3 remain unchanged when controlling for these other factors; specifically, deleterious rare variants show larger effects on the tail end of the IQ distribution in early childhood which attenuate with age, whereas the attenuation of the direct effect of PGI_EA_ is strongest in the top half of the IQ distribution These results indicate that the relative influences of factors that best predict which children will have cognitive difficulties or will excel cognitively across childhood differ from those that best predict average IQ.

## Discussion

Here we explored the contribution of common and rare variants to cognitive performance longitudinally across childhood and adolescence. We showed that common and rare variants differ in their patterns of effects on IQ and school achievement across time. While the effect of rare damaging variants attenuates between childhood and adolescence (Figure 2), the effect of common variants associated with educational attainment and cognitive performance in adulthood increases (Figure 1). In both cases, these changes over time are due to direct genetic effects, with no evidence for an age interaction effect of non-transmitted common or rare alleles in parents (Figure 1B), nor indeed for any effects of non-transmitted rare alleles on average IQ at all (Figure 2B). Our common variant findings accord with those of Malanchini *et al.* who showed that the effect of a polygenic score for non-cognitive skills on academic achievement increased over development in a different cohort^51^.

In theory, the increasing effect of polygenic indices for educational attainment and adult cognitive performance with age could simply be because the underlying SNP effects have been estimated on adult phenotypes which are better correlated with IQ in later childhood than early childhood^52,53^. Having said that, we observed suggestive evidence for an increase in the total SNP heritability of IQ with age in ALSPAC (p=0.053) (Table S2), consistent with the trend reported in another cohort^42^. Additionally, the common variant genetic correlations between IQ measured at the different ages and educational attainment measured in adulthood were not significantly different from 1 (Table S3), suggesting the increase in PGI effects are most likely due to genome-wide amplification of genetic effects as opposed to different common variants impacting cognitive ability differentially at different ages^11^. By contrast, our finding that the effects of rare variants attenuate as children age cannot be explained by the way the variant weights were obtained; this is because the weights are based on estimates of evolutionary negative selection against deleterious variants in each gene in large population-based cohorts of adults^46^, rather than through genotype-phenotype association studies.

Genetic studies typically test for genetic effects on the mean of the phenotype, although some have also considered effects on variance^54–58^ or differential effects on different quantiles of anthropometric traits and biochemical measures^59^. Here, we examined the longitudinal effects of common and rare variants on different quantiles of the IQ distribution, which essentially tests for an effect on the phenotypic variance and skew. We found that the main effects of polygenic scores and of RVB_Missense_ did not differ by quantile, whereas the main effect of RVB_pLoF_ was significantly stronger at the 5th than 95th percentile (Figure 3). However, the increasing effects of PGI_EA_ and PGI_Cog_ on mean IQ with age were strongest for the top half of the phenotype distribution, whereas the attenuating effects of RVB_pLoF_ were strongest in the lower tail (Figure 3). The latter observation appears to be driven by RVB_pLoF_ having a larger effect on the 5th than the 95th percentile of IQ at age 4 and 8, but similar effects at the distribution tails by age 16 (Figure S4). Results based on measures of cognitive performance in MCS and UK Biobank were concordant with these observations from ALSPAC (Extended Data Figure 7B). These results imply that the genetic profile of children in the highest and lowest ranges of cognitive ability differ across development (Extended Data Figure 9). Figure 5 summarizes our key findings for the IQ trajectories of individuals with different common and rare variant genetic backgrounds; children in the high versus low PGI_EA_ groups become more stratified across development, with more of the stratification occuring due to increasing IQ in the high PGI_EA_ group, while the stratification in IQ due to RVB_pLoF_ is strongest at the earlier ages and attenuates particularly in the high RVB_pLoF_ group.

**Figure 5.**
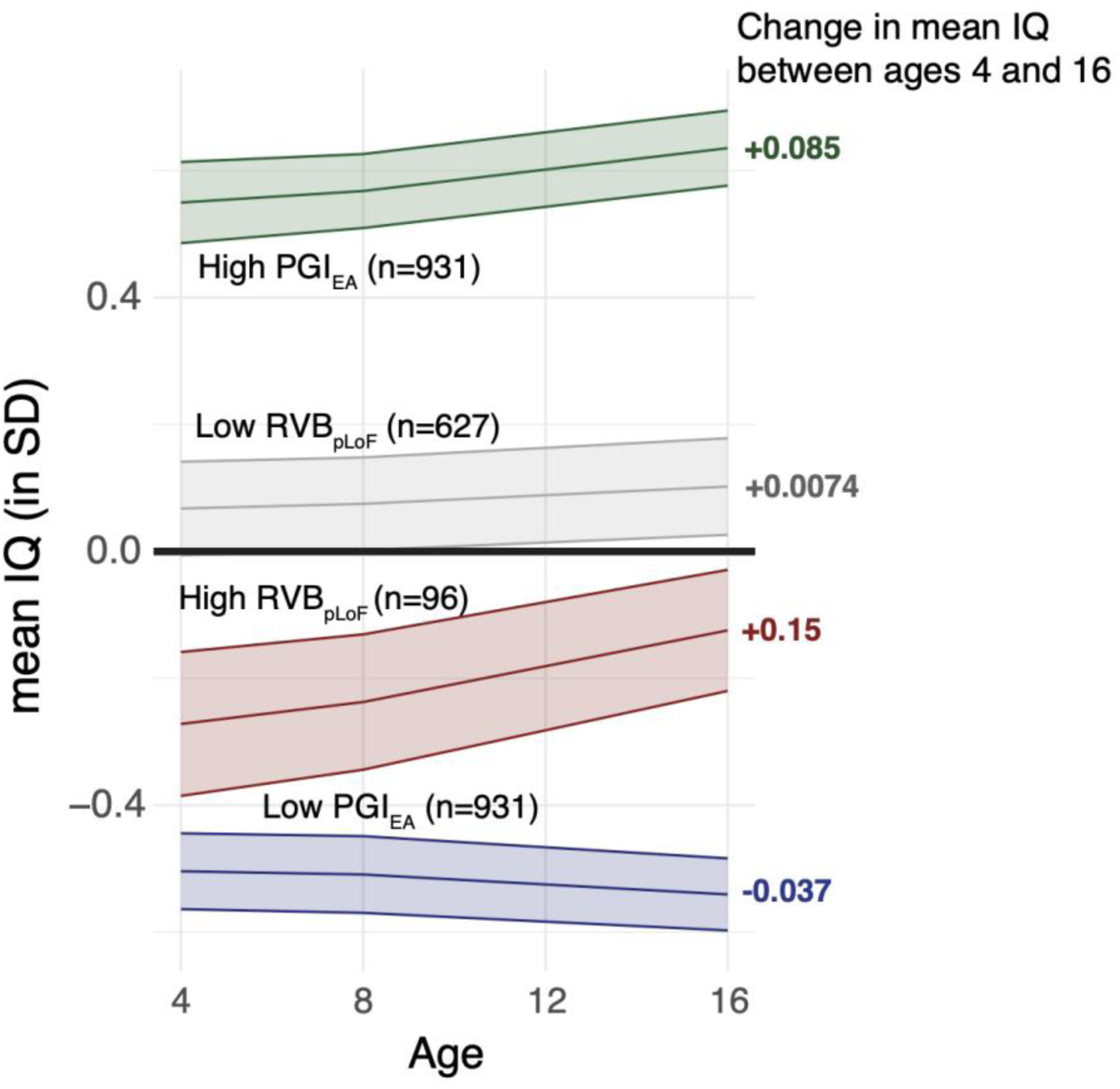
Summary of phenotypic trajectories across development, as inferred in this work. The plot shows average trajectories of IQ (inferred post-imputation) for individuals grouped by genetic measures. The high/low PGI_EA g_roups are individuals with a value that is one or more standard deviations above/below 0. The low RVB_pLoF g_roup are individuals with RVB_pLoF b_elow 0.01 (approximately 9.6% of individuals) and the high RVB_pLoF g_roup are individuals with RVB_pLoF g_reater than 0.4 (approximately 1.5% of individuals); these cutoffs were chosen arbitrarily for illustrative purposes. The change in mean genetic measure/IQ between ages 4 and 16 are shown on the right hand side of the plots. Bands indicate 95% CI. The decrease in mean IQ in the low PGI_EA g_roup is explained by the fact that, due to PGI_EA e_ffects becoming stronger towards the top of the IQ distribution, those that had a high IQ earlier in development will tend towards their genetically predicted lower IQ at later ages, thereby lowering the average IQ in this group.

Our results imply that there is higher variance in IQ in early childhood amongst individuals with higher RVB_pLoF_, which could be potentially due to gene-by-environment or gene-by-gene effects, or simply to stochasticity. Due to this higher variance, there are more people who cross the diagnostic threshold for intellectual disability than there are amongst those with low RVB_pLoF_, but there are also relatively more people within the typical IQ range, which could explain why these variants have variable and reduced penetrance. Multiple neurodevelopmental conditions exhibit a high degree of phenotypic variability, even amongst carriers of the same causal mutation, such as in neurofibromatosis type 1^60^. Although quantitative measures of intellectual impairment are relatively rare in studies of these conditions, there is evidence, for example, that patients with tuberous sclerosis complex (TSC) have not only lower *mean* IQ but also higher *variance* in IQ than unaffected sibling controls (F-test for a difference in variance between cases and controls; by our analysis, p=0.004 for TSC1, p=0.033 for TSC2)^61^. Taken together, our results support two hypotheses for the incomplete/variable penetrance of rare NDC-associated variants. Firstly, these variants impact variance in cognitive ability early in life, consistent with heterogeneous phenotypes of children with NDCs. Secondly, they have larger effects on average cognitive ability in early childhood than later in life, consistent with apparently clinically unaffected parents passing on these pathogenic variants to affected children.

The increasing heritability of IQ with age is well established but its causes remain unclear^11^. In contrast to our results for rare variants, we found evidence across cohorts for increasing *direct* genetic effects of PGIs with age, particularly at the top of the IQ distribution; to our knowledge, this has not been previously demonstrated. Recent work by Morris et al. using longitudinal twin modeling^62^ found evidence that the increase in heritability of cognitive performance across development is due to pervasive genetic innovation (i.e., new genetic effects coming into play across time, resulting in genetic correlations between ages that are less than one). In contrast, our molecular genetic results based on common variants suggest that amplification (i.e., identical genetic effects increasing proportionally with age) is the primary process. However, it is likely that these heterogeneous effects across the IQ spectrum that we have observed violate the assumptions of the twin model of Morris et al., and may lead to the appearance of genetic innovation where none exists. The increasing direct effect of common variants on IQ may be relevant to the so-called ‘Matthew effect’, the phenomenon whereby individual differences in ability compound over time and increase gaps in cognitive and academic outcomes as children age^63–66^. If this effect is particularly driven by those with high cognitive performance, this could explain the increase in common variant effects over time that we observe for the upper tail of the phenotype distribution. Several potential mechanisms could drive this, including evocative gene-environment correlations, such as high-performing children being selected into more from cognitively-stimulating environments, or active gene-environment correlations, such as high-performing children being more effective learners and hence improving their cognitive performance further relative to their peers^67^.

When comparing the relative magnitude of the main effects of genetic versus other factors on IQ in ALSPAC, we found that the RVB contribution was slightly larger than but broadly comparable to those of the perinatal exposures, and the direct effect of common variants associated with EA was comparable to that of parental EA (Figure 4A top). This highlights the important contribution of genetic variation across the allele frequency spectrum to variance in cognitive performance. We showed that inherited rare damaging coding variants explain more variance in childhood IQ in the general population than do *de novo* mutations (Supplementary Note 4), in contrast to what is observed in cohorts of children with neurodevelopmental disorders^22,33^. We further showed that the effects of both rare damaging variants and perinatal factors on cognitive ability attenuate as children age (Figure 4A bottom). This may be due to acute, time-limited effects during early neurodevelopment that can be attenuated by later plasticity, potentially in response to environmental influences. If this is true, one might predict that the attenuation of these effects with age are strongest in children from less deprived households. Testing this hypothesis will need larger datasets than studied here. In contrast to these other factors, the effect of parental EA on average IQ does not change across development, whereas the direct effect of EA-associated common variants increases (Figure 4A bottom), suggesting a cumulative influence. These observations have important implications for how we identify people at risk of poor outcomes in early versus in later life and highlight the value of considering genetic data in studies of social and environmental factors influencing cognitive outcomes.

There are several limitations of this study. Firstly, our primary findings about changing genetic effects with time are based on measurements of IQ at only three time points in a single cohort (ALSPAC). There is a dearth of cohorts with longitudinal measures of IQ in childhood and adolescence with sufficient sample size and appropriate genetic data for us to attempt direct replication. Thus, we have relied on replication with school grades measured at two timepoints in ALSPAC (not totally independent of our IQ results, given the overlapping sample) (Extended Data Figures 2 and 4; Figure S5), or, for some analyses, a single measure of cognitive performance from MCS and UK Biobank (Extended Data Figures 7 and 10, Figures S1 and S6), which we found to show strong genetic correlations with IQ in ALSPAC (r_g_ 1.09, 95% CI [0.97-1.21]) and with adult cognitive performance^44^ (r_g_ 0.89, [0.69-1.09]) and EA^47^ (r_g_ 0.89, [0.72-1.08]). Broadly, the results from these replication analyses supported our primary findings in ALSPAC. Secondly, IQ tests are less reliable in early life^68^, and those used in ALSPAC may be less accurate at measuring the tails of the distribution^69^. If the accuracy at the tails differed between the tests used at different ages, this may have induced spurious interaction effects between the genetic scores and age. However, this does not seem a likely scenario given that we see age interaction effects in opposite directions for common versus rare variants for the same trait, and replicate trends in other cohorts and with other cognitive performance measures. Thirdly, the genetic scores we have used to capture the effects of common and rare variants under-estimate the total liability conferred by these variants. Thus, the questions we have addressed here should be revisited in future once much larger common and rare variant association studies are available, including using whole-genome sequence data to capture the effects of noncoding variants.. A final limitation is that our results may be affected by non-random missingness within the cohorts and biased ascertainment and attrition across time, known to be present in both birth cohorts^37,70^. We have attempted to mitigate non-random missingness by imputation of missing IQ measures in ALSPAC (Figure S1) and by Mendelian imputation of unmeasured parental genotypes for our trio-based PGI analyses. To mitigate ascertainment bias and attrition, we incorporated weights in MCS to render the sample representative of the whole UK population. In general, our conclusions in ALSPAC were unchanged when analyzing IQ before versus after imputation, although the latter was obviously better powered, and there were some differences particularly in the findings from the quantile regressions (Supplementary Note 7; Figure 3BC *versus* Figure S3; Figure S4A *versus* S4B). One analysis that is likely to have been impacted by ascertainment bias is the estimate of the fraction of the rare variant effect on IQ that is due to *de novo* mutations, which we have probably over-estimated, as noted in Supplementary Note 4 (Extended Data Figure 6).

Our study prompts several strands of future work. Perhaps most importantly, future research should extend upon our findings to investigate how genetic and other factors might be best employed to identify children at risk of poor cognitive outcomes, so that interventions may be targeted at those who most need help. It should also seek to confirm the relevance of our observations to the incomplete penetrance of rare damaging variants in NDCs. If we had NDC cohorts with genetic and longitudinal phenotype data from both affected children and parents who appear to be clinically unaffected, we could directly test whether in fact, these variants were associated with reduced cognitive ability or educational achievement earlier in the parents’ lives, and whether their effects on these traits attenuated as they aged. Finally, future work should seek to better understand why there are differential time-varying genetic effects on children depending on their level of cognitive ability, and why the effect of our rare variant burden scores attenuates with time. For example, is the latter observation purely driven by the expression patterns of evolutionarily constrained genes that are more heavily weighted in this score (Extended Data Figure 5A), with these prenatally-expressed genes becoming progressively less important in brain function as children age? Or is it that children with particularly marked cognitive deficits in early childhood, who appear to be particularly influenced by these rare variants (Figure 3C), are targeted for interventions which help to mitigate the effects of these rare variants over time? More broadly, our results suggest that heteroscedastic and time-varying effects of genetic variants on human phenotypes deserve more exploration.

## Abbreviations

GWAS: Genome-wide association study;
NDCs: neurodevelopmental conditions;
SNP: single nucleotide polymorphism;
ALSPAC: Avon Longitudinal Study of Parents and Children;
MCS: Millennium Cohort Study;
IQ: Intelligence quotient;
PGI: polygenic index;
EA: educational attainment;
Cog: the cognitive component of educational attainment;
NonCog: the non-cognitive component of educational attainment;
RBV: rare variant burden;
pLoF: predicted loss of function;
TSC: tuberous sclerosis complex;
QC: quality control;
PCA: principal component analysis;
MAF: minor allele frequency;
HWE: Hardy-Weinberg Equilibrium;
LD: linkage disequilibrium.

## Acknowledgements

We thank the Human Genetic Informatics group at the Wellcome Sanger Institute for assisting in the generation and preparation of the whole exome sequencing data for the ALSPAC and MCS cohorts and general technical support. We thank Alexandra Havdhal for helpful discussions.

ALSPAC: We are extremely grateful to all the families who took part in ALSPAC, the midwives for their help in recruiting them, and the whole ALSPAC team, which includes interviewers, computer and laboratory technicians, clerical workers, research scientists, volunteers, managers, receptionists and nurses. The UK Medical Research Council and Wellcome (Grant ref: 217065/Z/19/Z) and the University of Bristol provide core support for ALSPAC. This publication is the work of the authors and Hilary Martin will serve as a guarantor for the contents of this paper. Genome-wide genotyping data was generated by Sample Logistics and Genotyping Facilities at the Wellcome Sanger Institute and LabCorp (Laboratory Corporation of America) using support from 23andMe. This research was specifically funded by the UK Medical Research Council and Wellcome (Grant ref: 076467/Z/05/Z) and the Department for Education and Skills (Grant ref: EOR/SBU/2002/121). A comprehensive list of grants funding is available on the ALSPAC website: http://www.bristol.ac.uk/alspac/external/documents/grant-acknowledgements.pdf

MCS: We are grateful to the Centre for Longitudinal Studies (CLS), UCL Social Research Institute, for the use of these data and to the UK Data Service for making them available. However, neither CLS nor the UK Data Service bear any responsibility for the analysis or interpretation of these data.

UKB: This research has been conducted using the UK Biobank Resource under application number 44165.

This research was funded in part by Wellcome (grant no. 220540/Z/20/A, “Wellcome Sanger Institute Quinquennial Review 2021–2026”). For the purpose of open access, the authors have applied a CC-BY public copyright license to any author accepted manuscript version arising from this submission. D.S.M. is supported by a Gates Cambridge Scholarship (OPP1144).

## Author contributions

DSM conducted all of the analyses. DSM, MK, PD, QH, WH, and EW carried out data preparation and quality control with supervision by SL, MEH, and HCM. MK led the whole exome sequencing data preparation for ALSPAC and MCS with assistance from DSM and EW. PD conducted the *de novo* variant calling in ALSPAC and MCS and conducted variant quality control for the UK Biobank whole exome sequencing data. WH conducted sample quality control for the UK Biobank whole exome sequencing data and assisted with preparation of UK Biobank exome data. OW assisted with selection of other non-genetic factors and generated PGI SNP weights. DSM, OW, QH, RA, MEH and HCM provided key intellectual input. HCM supervised the analyses and directed the study. DSM and HCM wrote the first draft of the manuscript with input from MEH. All authors read and commented on the final manuscript.

## Declarations

MEH is a co-founder of, consultant to and holds shares in Congenica, a genetics diagnostic company. The remaining authors declare no competing interests.

## Data availability

Researchers can apply to access data from ALSPAC (https://www.bristol.ac.uk/alspac/researchers/access/), MCS (https://cls.ucl.ac.uk/dataaccess-training/data-access/), and UK Biobank (https://www.ukbiobank.ac.uk/enable-your-research/apply-for-access).

## Online Methods

### Cohorts

We report results from the Avon Longitudinal Study of Parents and Children (ALSPAC)^37,71^, Millennium Cohort Study^38^ (MCS), and UK Biobank cohorts.

### ALSPAC

In ALSPAC, pregnant women with expected deliveries between April 1st 1991 and December 31 December were recruited in the greater Bristol area (formerly Avon county), resulting in an initial sample of 14,541 pregnancies enrolled into the study, of which 13,988 resulted in live births of children surviving to age 1. Data collected after the age of 7 was available for an additional 906 pregnancies from other phases of enrollment, which resulted in an additional 913 children that survived to age 1. At initial enrollment, 14,203 unique mothers were in the study, which increased to 14,833 unique mothers enrolled after the additional phases of enrollment (G0 mothers). The partners of G0 mothers (G0 partners) were also invited to participate in the study, of which 12,113 provided data at one point in the study and 3,807 are currently enrolled. From birth to early adulthood, mother and children were followed up with questionnaires and clinical and psychometric data collection across several time points. Biosamples used for genotyping and exome sequencing were obtained from most children and some of the mothers and fathers.

In the current study, data from 6,495 unrelated children with European inferred genetic ancestry (G1 children; described below) were used, along with genetic and/or survey data collected on 4,968 G0 mothers and 4,563 G0 partners. Please note that the study website contains details of all the data that is available through a fully searchable data dictionary and variable search tool: http://www.bristol.ac.uk/alspac/researchers/our-data/. Ethical approval for the study was obtained from the ALSPAC Ethics and Law Committee and the Local Research Ethics Committees. Consent for biological samples has been collected in accordance with the Human Tissue Act (2004). Informed consent for the use of data collected via questionnaires and clinics was obtained from participants following the recommendations of the ALSPAC Ethics and Law Committee at the time. At age 18, study children were sent ’fair processing’ materials describing ALSPAC’s intended use of their health and administrative records and were given clear means to consent or object via a written form. Data were not extracted for participants who objected, or who were not sent fair processing materials.

### MCS

MCS recruited 18,552 pregnant mothers (2000-2002) using a sampling scheme to ensure a nationally representative sample across the UK, as previously described^38^. Mothers and children were followed longitudinally and had genetic data collected at age 14 from children, mother, and fathers where available. Ethical approval for the collection of saliva samples from these individuals as part of the sixth sweep was obtained from London-Central Research Ethics Committee. Genotype and newly generated exome data are available for ∼8,000 and ∼7,000 children, respectively (∼13,000 and ∼7,000 parents).

### UK Biobank

The UK Biobank is a prospective cohort of over 500,000 individuals sampled throughout the UK between 2006 and 2010. Individuals have extensive phenotype data and genetic data including genotype array ^72^ and whole exome sequence data^73^.

### Cognitive performance measures across cohorts

In this study we used various cognitive performance and school performance measures across the three cohorts.

In ALSPAC, children had IQ measured at ages 4 (Wechsler Preschool and Primary Scale of Intelligence), 8 (Wechsler Intelligence Scale for Children), and 16 (Wechsler Abbreviated Scale of Intelligence). Linked educational records are available, including national standardized exam scores in English, Math, and Science administered at the end of Key Stage 2 (Year 6 at around 11 years old) and Key Stage 3 (Year 9 at around 14 years old). To create a composite measure of academic performance, we standardized each score and, at a given Key Stage examination point, computed a one-factor model score using *factanal* function in R using the Bartlett method for scoring, explaining 74% and 77% of the variance at Key Stage 2 and 3 exams, respectively. The factor loadings and factor variance explained for the three exams were nearly identical at Key Stage 2 and 3 (Table S1), implying longitudinal invariance.

In MCS, children completed various cognitive performance tests at several ages. Those at later ages had a greatly reduced sample size, so we focused on those at ages 3 to 7. These included Bracken School Readiness at age 3, reading vocabulary at ages 3 and 5, pattern construction at ages 5 and 7, and word reading and progress in math at age 7. These were summarized into a single cognitive performance measure using a one-factor model score using the *factanal* function in R and Bartlett method for scoring, explaining 39% of the variance.

In UK Biobank, individuals were asked thirteen questions testing verbal-numerical reasoning in a limited time frame during the initial assessment visit. We used the sum of questions answered correctly as a measure of fluid intelligence (data field 20016.0), as has been used previously in genetic studies of cognitive ability ^74^.

### Imputation of IQ values in ALSPAC

Imputation of missing IQ values was carried out using SoftImpute^75^ using rank.max = 4.5 and lambda = 4.5. Before imputation, all variables were standardized to have mean 0 and variance 1. To assess the imputation accuracy obtained using three potential sets of variables (described below), we set 100 random individuals with measured IQ values at a given age to missing, conducted phenotype imputation, and calculated correlations between the true measured values and the imputed values, repeating this procedure 100 times.

We considered the following sets of variables:

1) Base set: sex (kz021), birth weight (kz030), maternal and paternal socio-economic groups (b_seg_m and b_seg_p), and IQ values at ages 4 (cf813), 8 (f8ws112), and 16 (fh6280).
2) Expanded set: base set plus development score at age 2 (cf783), sociability score at age 3 (kg623a/c), additional verbal and performance IQ at age 4 (cf811, cf812), nonword repetition and multisyllabic word repetition at age 5 (cf470, cf480), communication score at age 6 (kq517), Skuse social cognition score at age 7 (kr554a/b), cognitive scores at age 8 including Sky Search, Diagnostic Analysis of Nonverbal Accuracy, nonword repetition, verbal and performance IQ, children communication checklist score (f8at062, f8dv443, f8dv444, 8dv445, f8dv446, f8sl100, f8sl101, f8sl102, f8ws110, and f8ws111, ku506a).
3) Auxiliary set: expanded set excluding base set variables.

When calculating the imputation accuracy using the expanded or auxiliary sets, we removed verbal and performance IQ at each age, although in practice very few children had only one measured at a given age.

For the imputation of the final IQ values, we used the expanded set and required that each individual had nonmissing values for at least one IQ test and at least 20% of the variables used for imputation. Final IQ values were standardized to mean 0 and standard deviation of 1.

### Genotype data preparation and imputation

#### ALSPAC

ALSPAC genotype data were generated and processed as described in ^71^. We further removed individuals with high genotype missingness >3%. We restricted analyses to autosomal SNPs with MAF >0.5%, missingness rate <3%, and that had HWE test p>1×10^-5^.

We used KING^76^ for relatedness inference and removed 152 samples who had first degree relationships but were reported as coming from different families and another 16 samples who did match available exome sequencing data supposedly for the same individuals resulting in 8,831 children, 9,302 mothers, and 1,706 fathers. To identify a set of unrelated children, we iteratively identified the child with the greatest number of genetically inferred relatives (3rd degree or closer) and removed them, recalculated the number of inferred relatives per child, and repeated the first step until no children were inferred to have any relatives. This resulted in a total of 6,495 unrelated children with genotype array and whole exome sequence data.

To identify individuals of genetically inferred European ancestry, we projected samples onto 1,000 Genomes phase 3 individuals^77^ using the smartpca function from EIGENSOFT version 7.2.1^78^. We used linkage disequilibrium (LD)-pruned SNPs (pairwise r^2^ <0.2 in batches of 50 SNPs with sliding windows of 5) with MAF > 5% and removed 24 regions with high or long-range LD, including the HLA^79^. All ALSPAC samples projected onto European ancestry samples. We then performed PCA identically on the unrelated ALSPAC samples and projected all individuals into the PCA space.

Prior to imputation, we removed palindromic SNPs, SNPs that were not in the imputation reference panel, and SNPs with mismatched alleles. We imputed the samples to the TOPMed r2 reference panel using the TOPMed imputation server^80–82^. We kept well imputed common variants with Minimac4 R^2^ >0.8 and MAF >1%.

#### MCS

MCS genotyped data were generated and processed as described in ^83^. A set of unrelated individuals with genetically-inferred European ancestry were described in a similar manner to ALSPAC, as described previously ^26^. We kept well imputed common variants with Minimac4 R^2^ >0.8 and MAF >1%.

### UK Biobank

UK Biobank genotype data were generated, processed, and imputed by UKB as described in ^72^. Sample quality control consisted of excluding individuals with >3% missingness, inconsistent sex, sex aneuploidy, or withdrawn consent and relatedness was similarly calculated using KING as previously described. As in the other cohorts, we kept autosomal SNPs with MAF >1%, missingness rate <3%, and passed the HWE test (p-value>1×10^-5^). To identify UKB individuals with genetically inferred European ancestry, we similarly projected samples onto the 1,000 Genomes Phase 3 individuals and assigned individuals to a genetic ancestry based on their Mahalanobis distance to the nearest continental ancestry group centroid using the top 6 PCs. Those more than 6 standard deviations from the centroid along any axis were removed. Unrelated individuals were identified using the iterative exclusion procedure as previously described, resulting in 387,531 unrelated individuals. PCs were recalculated in this subset of unrelated individuals using smartpca as previously described, and related individuals were projected into this PCA space.

### Calculating polygenic scores

SNP weights for polygenic scores were estimated using LDpred2-auto^84^, which does not require a tuning dataset and automatically estimates the required hyper-parameters from the discovery sample^85^. The LD reference panel was computed from unrelated individuals from the target dataset for a set of 1,444,196 HapMap3+^86^ variants^72^. GWAS summary statistics for EA^16^ excluding 23andMe or 23andMe only (used for PGI_EA_ UK Biobank samples), EA-Cog and EA-NonCog^44^ were matched with the list of overlapping SNPs.

Once the weights were generated, we scored individuals using the --score function in PLINK v1.9, which calculates the weighted sum of genotypes across a set of SNPs for each individual.

### Mendelian imputation of parental genotypes

When an individual had at least one parent with genotype data in the cohort, we imputed the expected parental genotype of the other parent. To do so, we used *snipar*^20^ across the three cohorts. We supplied relatedness inference using KING output and used the default options throughout. We used the previous weights derived from each GWAS using LDpred2-auto to calculate the PGIs in the full trios using the pgs.py script provided in the *snipar* package.

### GWAS of IQ and cognitive performance

We used the linear regression function *lm* in R to conduct GWASs on the IQ measures pre- and post-imputation in ALSPAC and the cognitive performance measure in MCS on unrelated genetically inferred European ancestry individuals, controlling for 10 genetic PCs and sex. We removed variants with MAF<1% or missingness >2%.

### Heritability and genetic correlations

We used both GREML-LDMS^41^ and LD score regression^87,88^ to estimate SNP heritabilities and genetic correlations from the summary statistics of the GWASs conducted on the IQ measures and external GWASs using the previously described LD reference panel on HapMap3 SNPs.

### Other exposures impacting IQ in ALSPAC and MCS

We considered the following exposures: maternal and paternal educational attainment (EA), weeks born preterm, and a composite variable representing maternal illness in ALSPAC only. For educational attainment, in ALSPAC we used the highest qualification reported by the parent and coded it as done previously reported^16^, with a university degree equalling 20 years of education, A levels as 13, O levels/vocational as 10, and Certificate of Secondary Education/no degree as 5 (c645a and c666a for the mother and father, respectively) and in MCS we used the equivalent self-reported derived variables in sweep 1 from each parent questionnaire (APLFTE00). Maternal illness was defined as a binary variable indicating whether or not a mother had at least one of the following conditions reported during the pregnancy in her obstetric clinical records: preeclampsia, anemia (DELP_1060), any diabetes (pregnancy_diabetes), or genital herpes, gonorrhea, syphilis, urinary tract infection, vaginal infection, or hepatitis B noted during delivery (DEL_P1050-1055). Weeks born preterm was coded as 40 minus the gestational age at birth in weeks assessed in a formal pediatric assessment (DEL_B4401). All variables were standardized to have mean zero and standard deviation 1.

### Exome sequencing data preparation

Whole exome sequencing data quality control for ALSPAC and MCS cohorts was carried out by the Human Genetics Informatics team at the Sanger Institute as described in ^89^. Briefly, GATK v4.2 was used to call short variants (SNVs and indels) in 11,994 samples from ALSPAC and 11,916 samples from MCS. Sample quality-control (QC) measures were employed to remove outliers on several metrics (e.g., heterozygosity, variant counts) and likely sample mismatches. To identify low quality variants (variant QC), a random forest was trained on pre-defined truth sets in each cohort individually. The random forest filtering was then applied in combination with genotype-level and missingness filters to balance precision, recall, true and false positive rates, and synonymous transmission ratios. Specifically, SNVs were filtered (genotypes set to missing) if they had an allele depth (DP) < 5, a heterozygous allele balance ratio (AB) < 0.2, or a genotype quality (GQ) < 20 (ALSPAC) or < 15 (MCS); Indels were filtered using these thresholds: DP < 10, AB < 0.3, GQ <10 (ALSPAC) or GQ < 20 (MCS). The variants were excluded if they failed the random forest filtering or if the fraction of missing genotypes (missingness) exceeded 0.5. The final dataset included 8,436 children and 3,215 parents in ALSPAC and 7,667 children and 6,925 parents in MCS. Calling and QC of *de novo* mutations is described in the Supplementary Methods.

In UK Biobank, we performed quality control for whole exome sequencing data on 469,836 participants within the UKB research analysis platform. First, we split and left-aligned multi-allelic variants in the population-level Variant Call Format files into separate alleles using *bcftools norm*^90^. Next, we performed genotype-level filtering using bcftools filter separately for Single Nucleotide Variants (SNVs) and Insertions/Deletions (InDels) using a missingness-based approach. Specifically, SNV genotypes with a depth lower than 7 and genotype quality lower than 20, or InDel genotypes with a depth lower than 10 and genotype quality lower than 20, were set to missing. We further tested for an expected alternate allele contribution of 50% for heterozygous SNVs using a binomial test, and SNV genotypes with a binomial test p-value ≤ 0.0001 were set to missing. Finally, we recalculated the proportion of individuals with a missing genotype for each variant and excluded all variants with a missingness value greater than 50%.

### Classification of deleterious rare variants

All variants were annotated using the MANE transcript from Ensembl^91^. In MCS and ALSPAC, rare pLoFs were defined as pLoFs annotated as high confidence by LOFTEE^35^ that had CADD^92^ (if SNVs) > 25, were not located in the last exon or intron, and had a gnom-AD^35^ V3 allele frequency of <3×10^-5^ (up to ∼ 5 occurrences in gnomAD r3 genomes & ∼ 10x in exomes) and an in sample allele frequency of <0.1% among the unrelated set of children from that cohort^91^. Rare damaging missense variants were defined using identical allele frequency and CADD filters to pLoFs but were additionally required to have an MPC score^93^ ≥ 2. Rare synonymous variants were defined with identical allele frequency thresholds.

In UKB individuals with inferred European genetic ancestry, we defined rare variants as those with a within-sample allele frequency <0.001%. For pLoFs, we retained only those variants defined as high-confidence pLoFs by LOFTEE and CADD > 25 as in MCS and ALSPAC. For missense variants, we defined the damaging missense variants by including variants with REVEL > 0.5, AlphaMissense^94^ > 0.56, and MPC > 2.

### Calculating rare variant burden scores

We calculated RVB using the following formula:

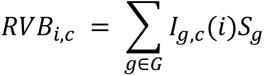

where *I*_*g*,*c*_(*i*) is an indicator function for whether an individual *i* has a variant of consequence *c* in gene *g*, *S*_*g* i_s the fitness cost for heterozygous carriers of a pLoF allele in gene *i* estimated in Sun *et al.*^46^, and *G* is the set of all autosomal genes.

For analyses to ascertain the relative contribution of *de novo* versus inherited variants (Supplementary Note 4), we also calculated a separate rare variant burden metric which we call constrained variant count:

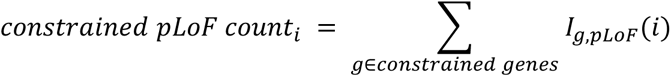

where *I*_*g,pLoF*_ (*i*) is an indicator function for whether an individual *i* has a pLoF in gene *g,* and *constrained genes* is the set of genes identified as constrained in ^46^.

### Sampling and non-response weights in MCS

Sampling weights were developed by MCS to adjust for the nonrandom sampling scheme devised for the study^95^. We used the full UK sampling weights. Nonresponse inverse probability weights were developed for each sample as described previously^26^. Briefly, we used logistic regression to predict whether an individual had complete data for a given regression. We then extracted the predicted values for all individuals with complete data from the logistic regression which correspond to their relative probabilities of having complete data, calculated the inverse of that value, and used that as the nonresponse weight. To combine sampling and nonresponse weights, we multiplied the two weights and incorporated them into the regressions.

### Associations between genetic measures and traits

Cross-sectional associations for a genetic score and IQ/cognitive performance measure were conducted using the *lm* function in R, restricting to unrelated samples with genetically-inferred European ancestry as follows:

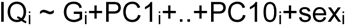

where G_i_ is the genetic score (PGI or RVB) for child *i* and PC1-10_i_ are their genetic PCs.

Genetic scores, unless otherwise stated, were standardized to mean 0 and standard deviation 1. In trio-based analyses, the parental genetic values were similarly standardized and regression was conducted as follows:

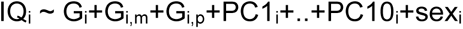

where G_i,m_ and G_i,p_ are the genetic scores for child *i*’s mother and father, respectively.

Mixed effect linear models were conducted using the *lme4* package in R^96^. The same covariates were used for the cross-sectional analysis with the addition of age, an age-by-genetic score interaction effect and a child-specific random intercept term as follows:

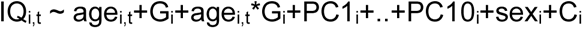

where age_i,t_ and IQ_i,t_ are child *i*’s age and IQ at age *t* and C_i_ is a child-specific random effect intercept. Age was coded as age-4 such that the genetic predictor intercept estimate was equivalent to the effect at the first age at which IQ data were collected. In the trio-based analyses, an age interaction effect was additionally modeled for the parental genetic values as follows:

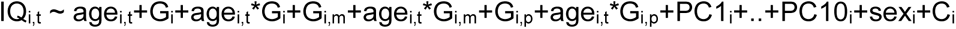

For cross-sectional and longitudinal quantile regression modeling, we used the R package *quantreg*^97^ and *rqpd*^98^, respectively, using the same models as above. Mixed-effects model standard errors were determined using bootstrapping with 500 bootstrap replications.

For the academic performance score, the cross-sectional models were performed in the same way described above, with the addition of age at testing in weeks as a covariate (ks2age_w and ks3age_w). As there were only two timepoints, a slightly different longitudinal model was used for the linear modeling (Extended Data Figures 2 and 4) as follows:

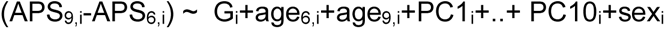

where APS_6/9,i_ and age_6/9,i_ are the academic performance scores and ages of individual *i* at Year 6 and 9, respectively. The coefficient estimated for G_i_ in this regression is equivalent to G’s effect at Year 9 minus that at Year 6 within an individual, i.e. the age interaction effect. To compare the effects for the quantile regression effect size estimates at Years 6 and 9, we conducted z tests between the effect sizes (Extended Data Figure 7A).

### Marginal and conditional associations between genetic and other factors and IQ

To assess the associations between genetic and other factors with IQ (Figure 4), we considered both marginal and conditional models. For marginal models, we conducted the following regressions:

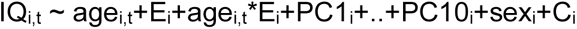

where E_i_ is the given measure of interest. In Figure 4, the coefficient on the E_i_ term (i.e. the main effect) is shown in the top panel, and the coefficient on the age_i,t_*E_i_ term (i.e. the interaction effect) in the bottom panel.

As maternal and paternal EA are highly correlated, we modified the model for the effects of parental EA as follows:

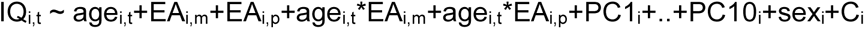

where EA_i,m_ and EA_i,p_ are the maternal and paternal EA respectively. Similarly, we fit the PGI_EA_ values for the child, mother, and father jointly in a trio model.

To calculate the variance explained by a given variable, we took the square of the standardized effect size of that variable’s main effect. To calculate the total variance explained by two uncorrelated variables (e.g. RVB_pLoF_ and RVB_Missense_ that have a correlation of -0.01, p=0.16), we summed the squares of their standardized effect sizes. To test for difference in effect size between the combination of RVB_Missense_ and RVB_pLoF_ *versus* other factors, we used the square root of the previous variance explained estimate as the effect size estimate and determined the standard error for that effect size by summing the square of the standard error for each effect estimate and then taking the square root of the new estimate. We were then able to use z tests to compare the effect size estimates as done previously.

For the full joint models, we included all genetic and other measures in a single model, with a main and age interaction effect for each measure. Since only a subset of probands had information on gestational age, we fitted the full joint model both with and without the ‘weeks born preterm’ variable.

## Extended Data Figures

**Extended Data Figure 1.**
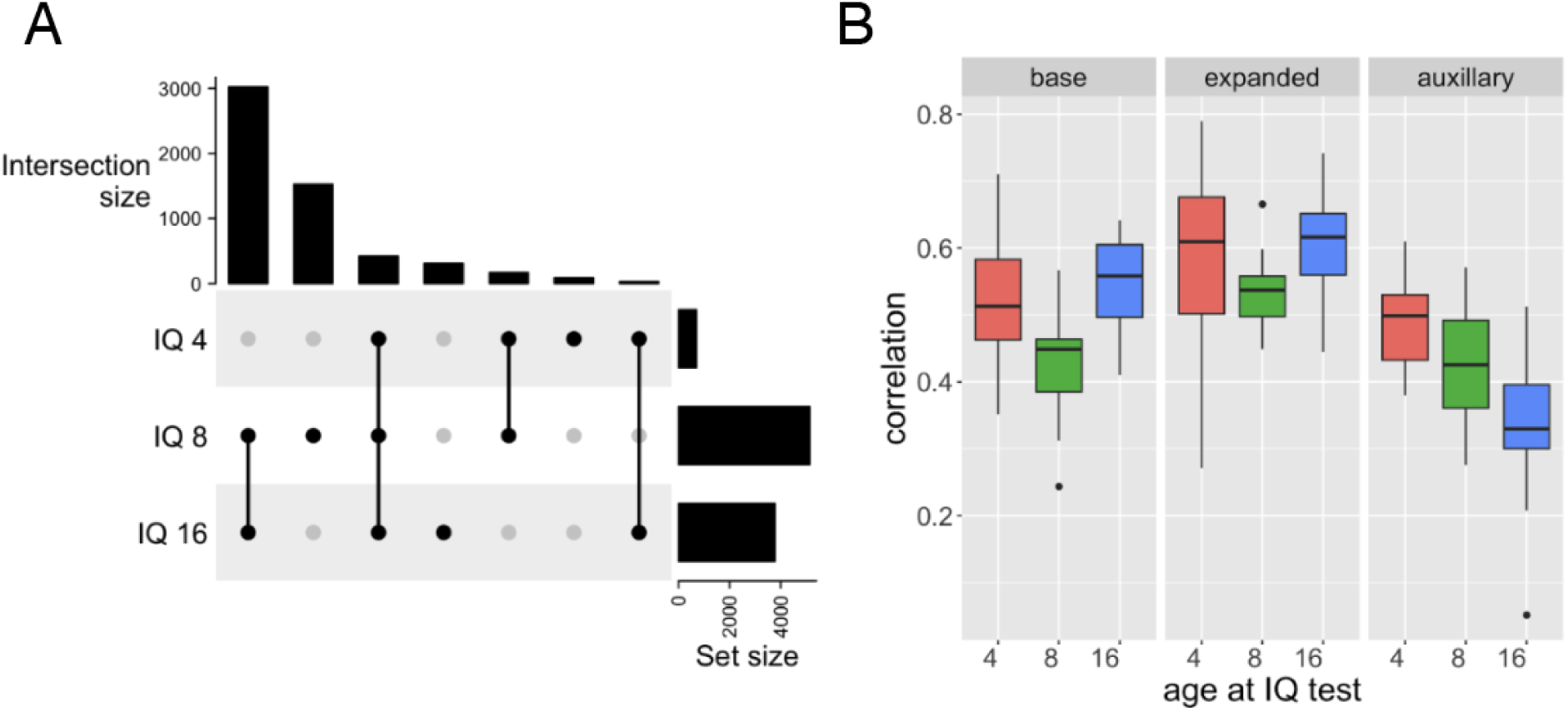
Assessing quality of IQ imputation across ages and sets of variables in ALSPAC. A) Upset plot indicating number of individuals with a given set of measured IQ values. B) Correlations between masked measured and imputed values for 100 random individuals for 100 trials for three different combinations of variables (base, expanded and auxiliary variables, as described in Methods) used in the SoftImpute imputation for IQ measured at age 4, 8, and 16.

**Extended Data Figure 2.**
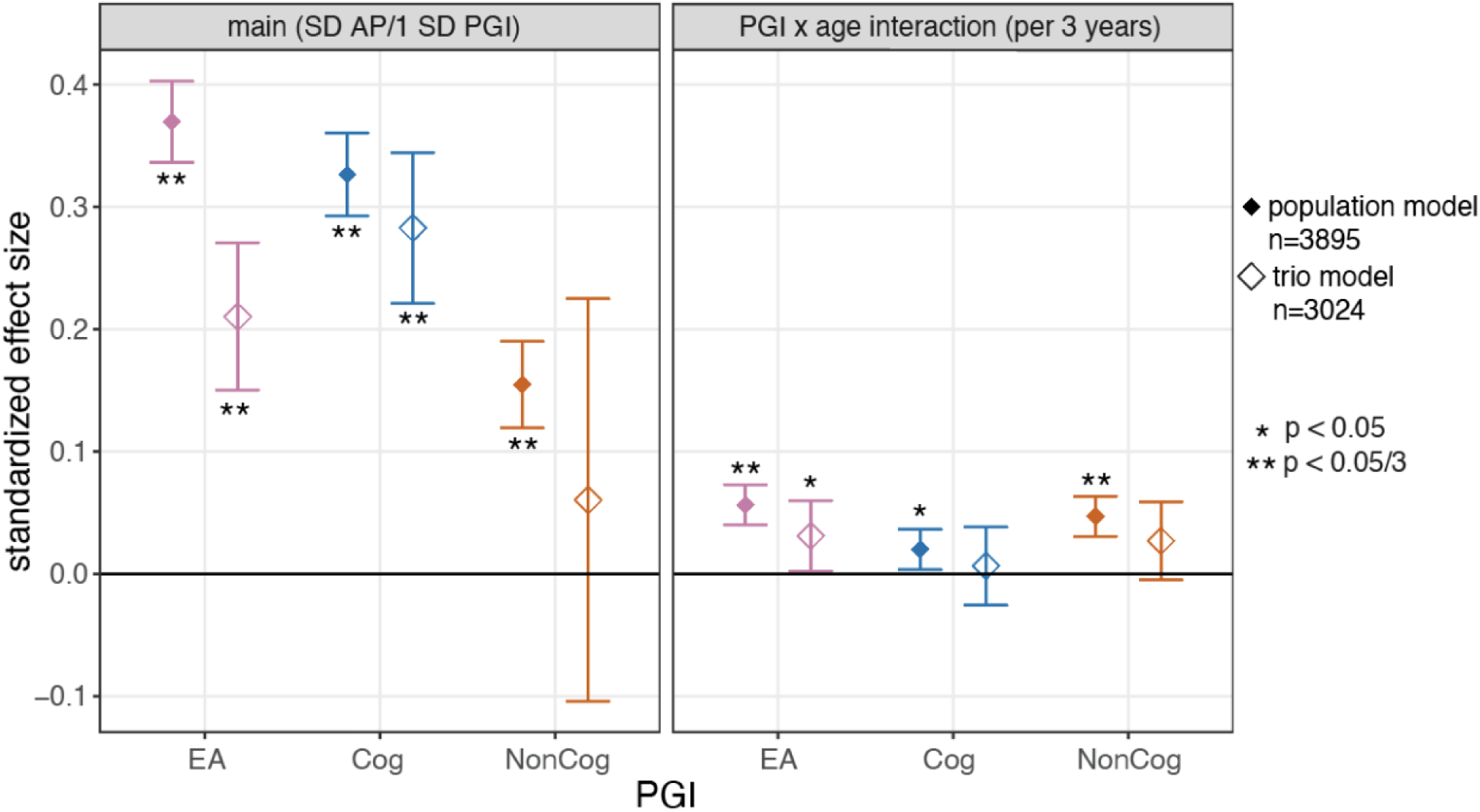
Association between PGIs and academic performance in ALSPAC. The points show the effect at age 11 (main effect, left panel) and the difference in effects between age 14 and 11 (PGI x age interaction, right panel) and error bars show 95% confidence intervals. Results are shown for the children’s PGIs not controlling (i.e. ‘population model’) and controlling (right) for parental PGIs. Population effect sizes are shown estimated in the full sample.

**Extended Data Figure 3.**
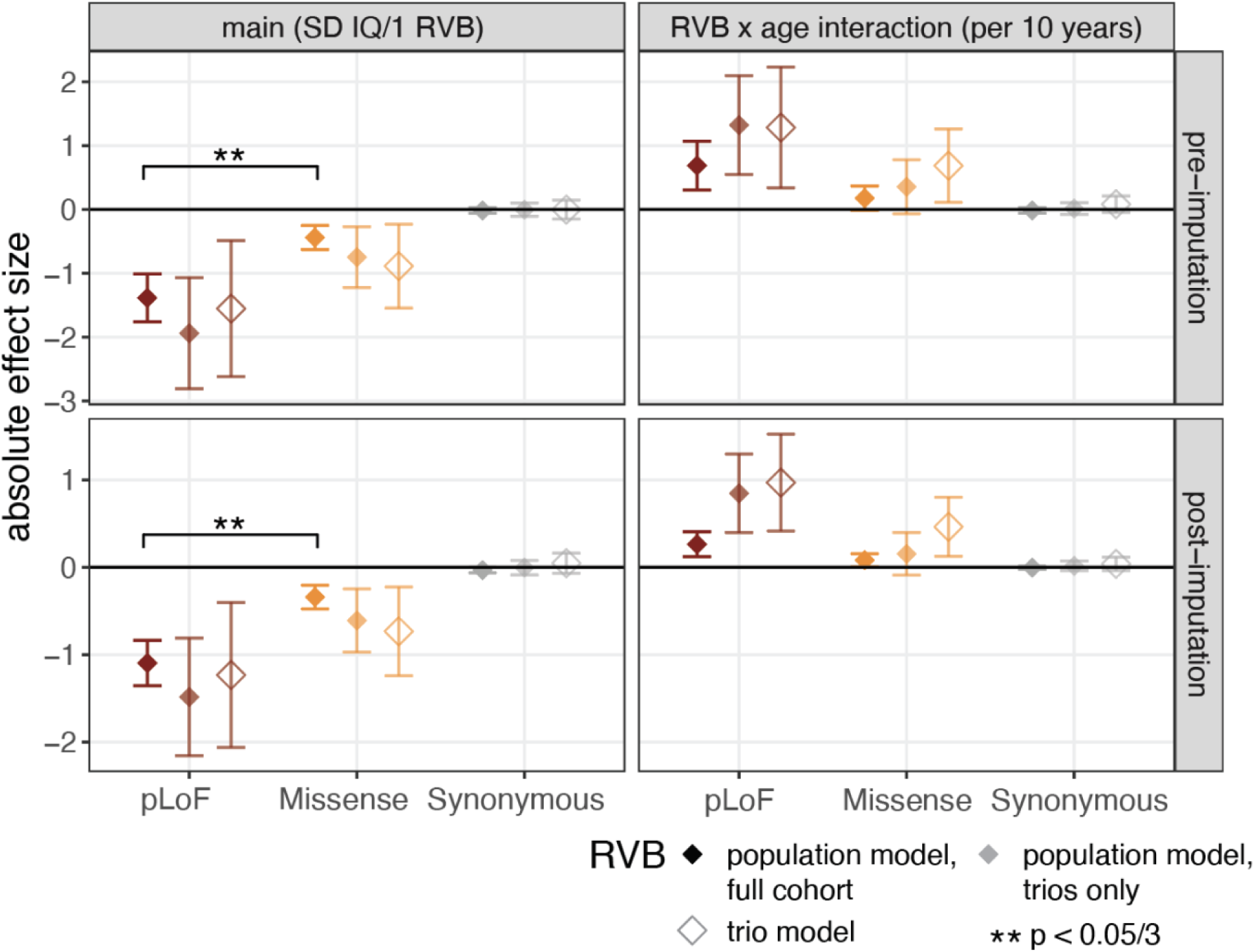
Association between unstandardized rare variant burden (RVB) and IQ across ages in ALSPAC, estimated with a mixed-effects linear model. (i.e. as for Figure 2 but with unstandardized rather than standardized RVB scores.) Absolute effects and 95% confidence intervals estimated for the main effects and RVB-by-age-interaction effects for RVB calculated with three different consequence classes (pLoF, missense or synonymous), either pre-(top) and post-imputation (bottom). Results for the children’s RVBs not controlling (i.e. ‘population model’) and controlling (right) for parental RVBs (i.e. trio model). Population effect sizes are shown estimated in the full sample (opaque) and in the subset of children with parental RVBs (translucent). The square brackets indicate significant comparisons highlighted in the text (z tests).

**Extended Data Figure 4.**
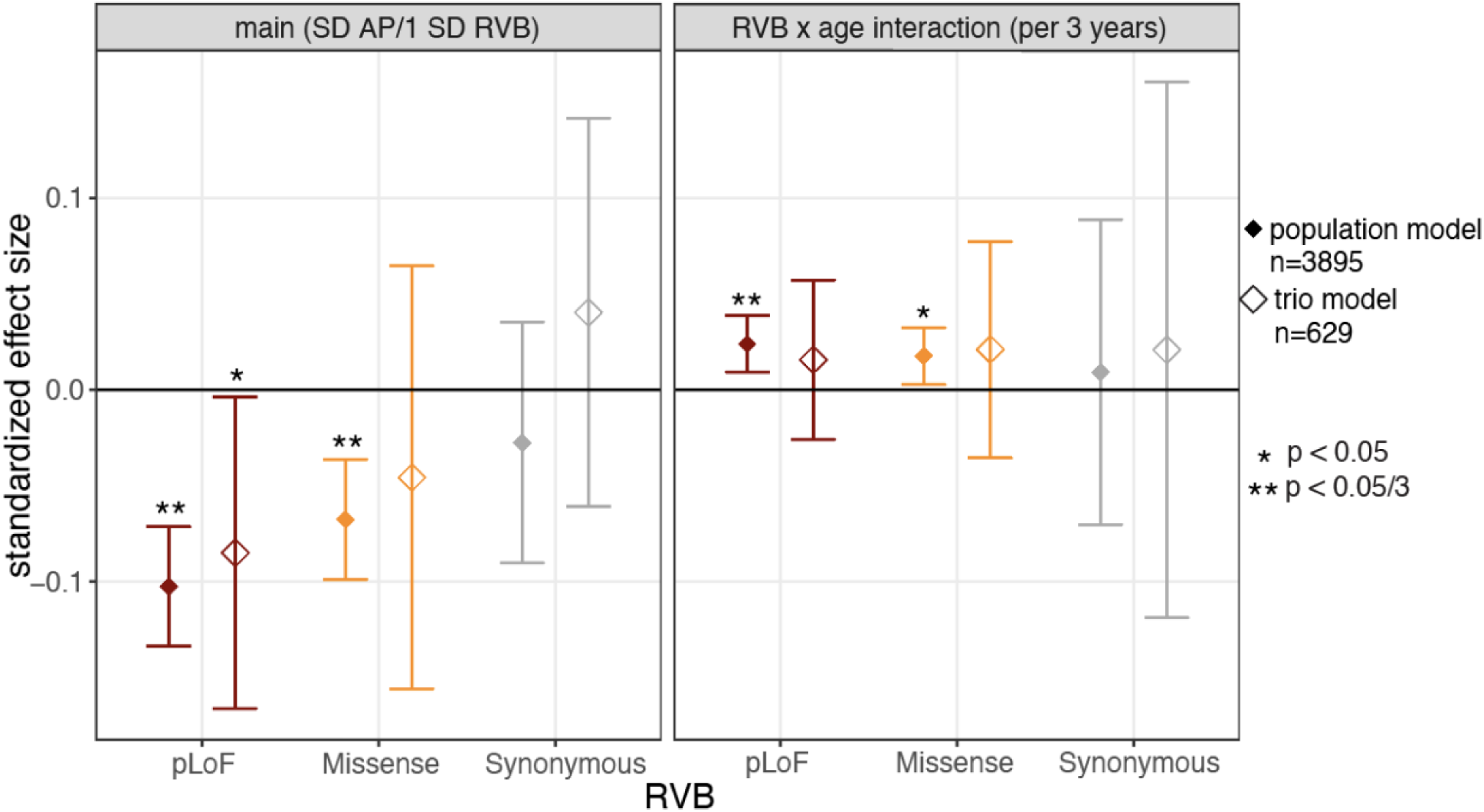
Association between RVBs and academic performance in ALSPAC. The points show the effect at age 11 (main effect, left panel) and the difference in effects between age 14 and 11 (RVB x age interaction, right panel) and error bars show 95% confidence intervals. Results are shown for the children’s RVB not controlling (i.e. ‘population model’) and controlling (right) for parental RVB. Population effect sizes are shown estimated in the full sample.

**Extended Data Figure 5.**
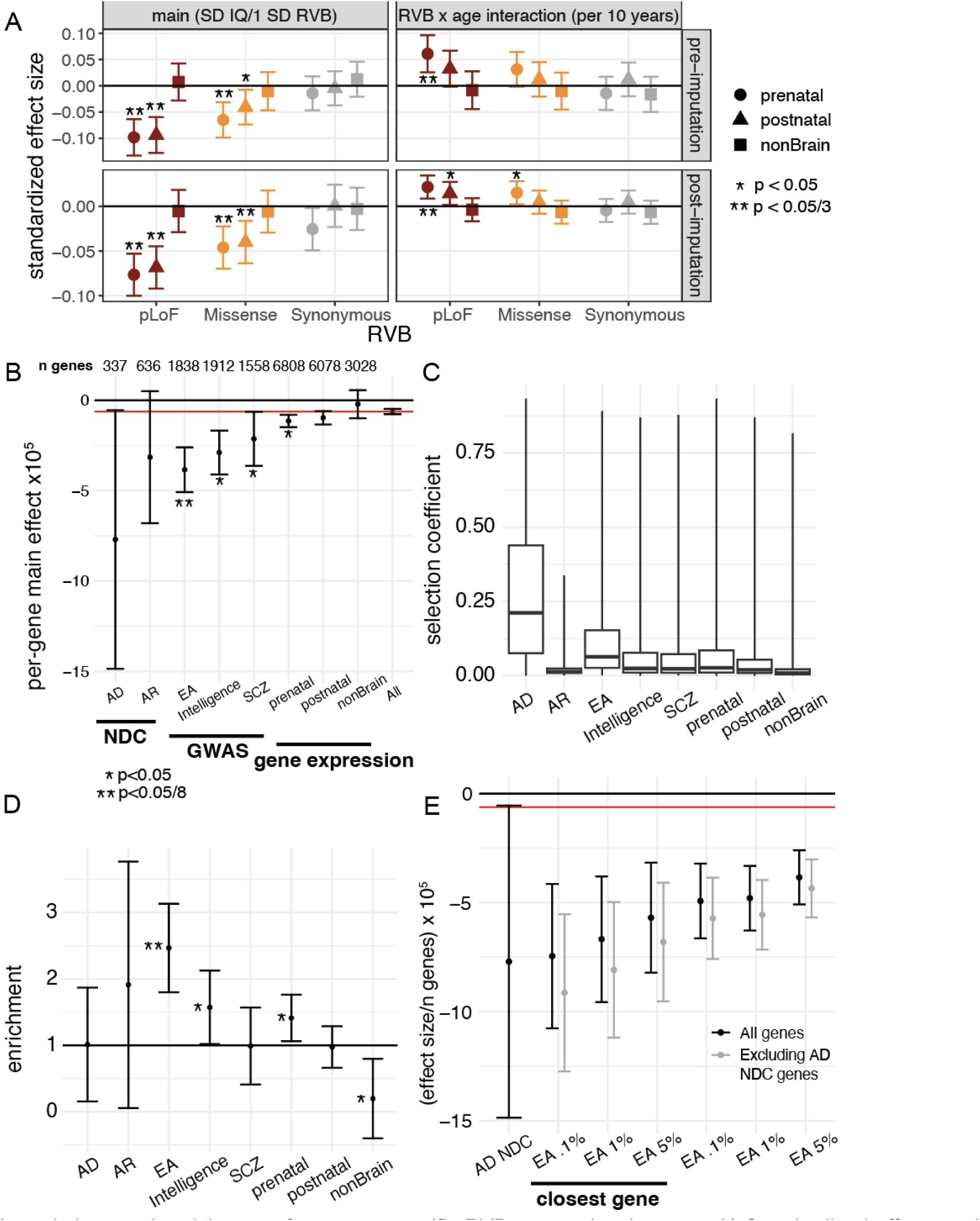
Associations and enrichment of gene set-specific RVB across development. A) Standardized effects and 95% confidence intervals estimated for main effects and RVB-by-age-interaction effects for each RVB in three mutually exclusive gene sets from Li et al^48^. These comprise genes in co-expressed clusters that are more highly expressed in prenatal or postnatal brain or that are not detected in the study (non-brain). B) Main effects of RVB_pLoF o_n IQ (post-imputation) stratified by gene set, divided by the number of genes in the given gene set, with 95% confidence intervals. Red horizontal line indicates the average effect for RVB_pLoF a_cross all genes. Asterisks indicate the p value for difference in per-gene effects between a given gene set and all genes using a z test, with * indication nominal significance and ** indicating bonferroni significance for 8 tests. C) Boxplots of the distribution of s_het (_selection coefficient against heterozygous pLoFs in that gene^46^) per gene set, where a coefficient of 0 indicates no selection against heterozygous pLoFs in a given gene and 1 indicates a 100% reduction in fitness for heterozygous pLoF carriers relative to non-carriers. D) Ratio of the main effect for RVB_pLoF f_or the indicated gene set relative to randomly sampled gene sets with matching underlying s_het d_istributions (enrichment). E) As in (B) for gene sets defined using different FDR threshold cutoffs based on gene prioritization in ^47^ (5%, 1%, 0.1%) and by restricting to prioritized genes at a given cutoff that are also the closest genes to the prioritizing SNP in the Lee et al. GWAS. Results are shown before (black) or after (grey) excluding overlapping genes from the set of autosomal dominant neurodevelopmental condition genes with a loss-of-function mechanism from DDG2P (AD NDC). AD/AR NDC: Autosomal dominant/recessive neurodevelopmental disorder genes with loss-of-function mechanism from DDG2P^99^, EA: educational attainment GWAS prioritized genes by Lee et al.^47^ at 5% FDR threshold, Intelligence: intelligence GWAS prioritized genes from Savage et al.^12^, SCZ: schizophrenia GWAS prioritized genes from Pardiñas et al.^100^

**Extended Data Figure 6.**
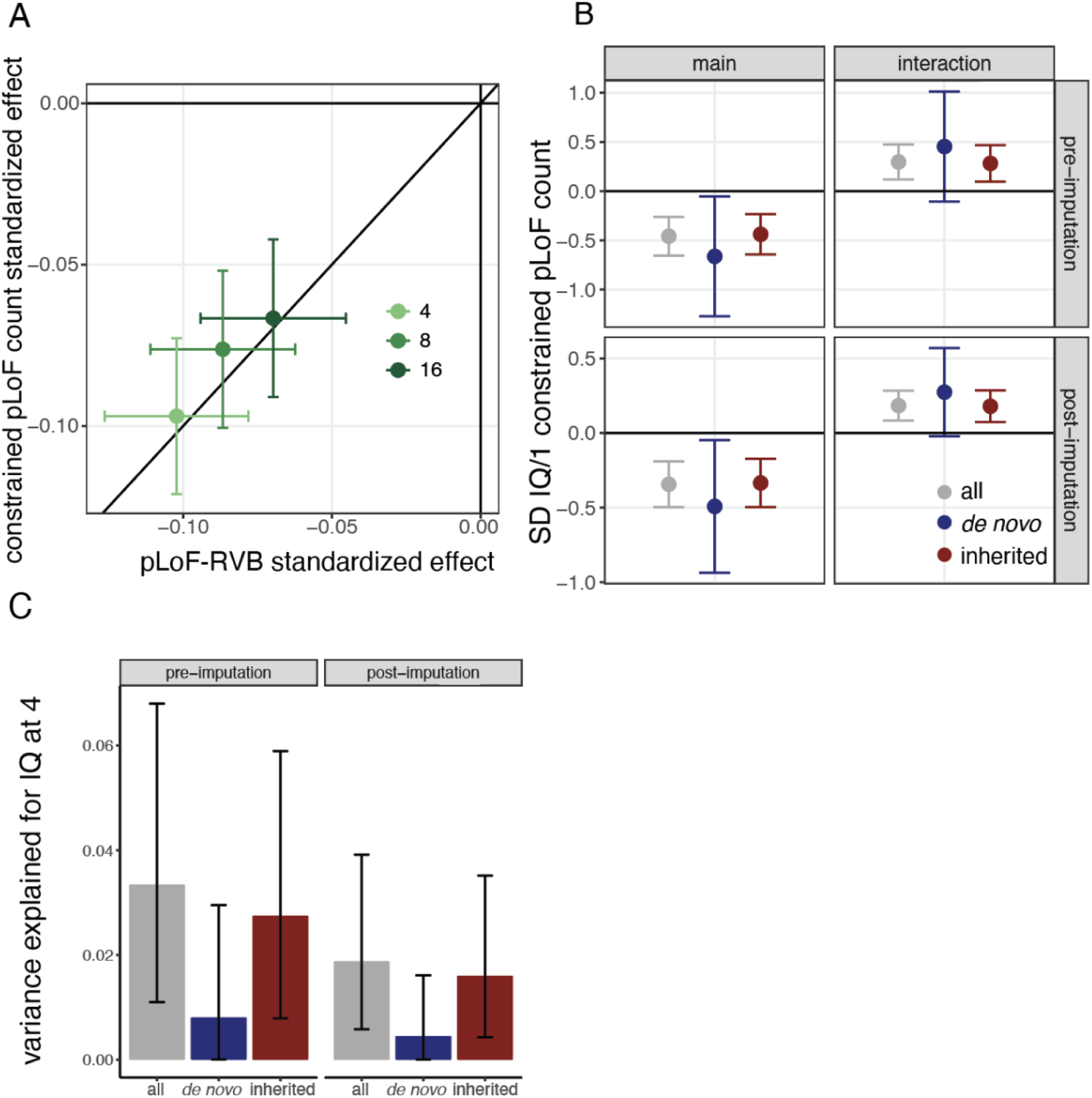
Association between inherited and *de novo* pLoF variant counts in constrained genes and IQ in ALSPAC. A) Standardized effects and 95% confidence intervals of RVB_pLoF a_nd constrained pLoF count (see **Supplementary Methods**) on IQ across ages post-imputation. Black line indicates the y=x line. B) Standardized effects for the main and RVB-by-age interaction effect from a longitudinal mixed-effects model of constrained pLoF counts on standardized IQ when considering all variants or *de novo* mutations and inherited variants separately, both pre- and post-imputation. C) Variance explained by constrained pLoF counts on standardized IQ when considering all variants or *de novo* and inherited separately, both pre- and post-imputation. See Figure S1 for results from a similar analysis of cognitive performance in MCS.

**Extended Data Figure 7.**
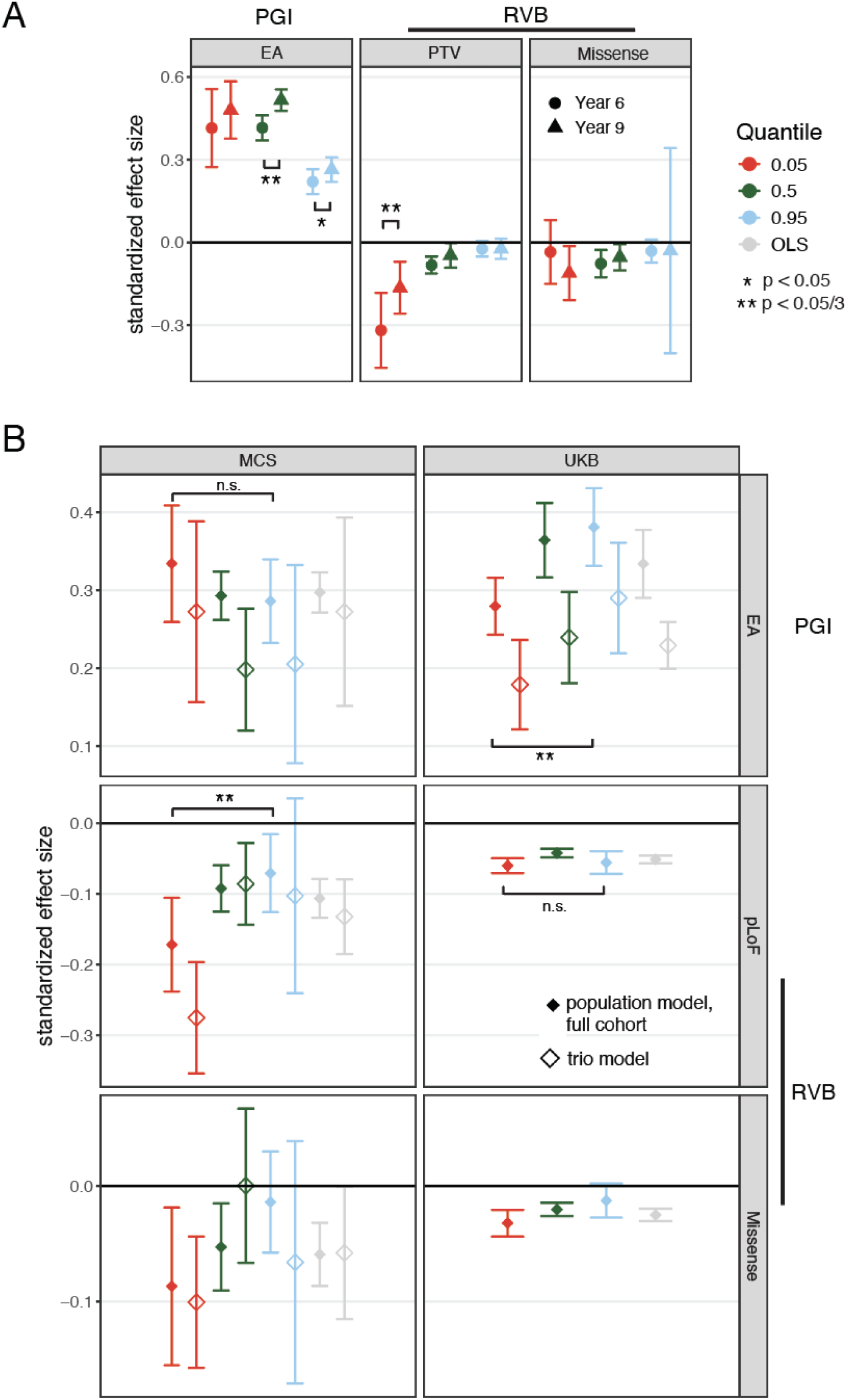
Influence of genetic measures on the tails of the phenotypic distribution using academic performance in ALSPAC and cognitive performance measures in MCS and UK Biobank. A) Standardized effects and 95% confidence intervals for quantile regression of the 5th (red), 50th (green), and 95th (blue) percentiles of genetic measures on academic performance in Year 6 and Year 9 in the full ALSPAC cohort showing population effects. The square brackets with asterisks indicate significant age interactions. B) Standardized effects and 95% confidence intervals for quantile regression of the 5th (red), 50th (green), and 95th (blue) percentiles and linear regression (OLS; gray) estimated from cross-sectional associations with cognitive performance measures in MCS and UK Biobank. The PGI_EA u_sed in UK Biobank was constructed from an EA GWAS in 23andMe only ^16^, as the GWAS excluding 23andMe includes UK Biobank. We only considered PGI_EA a_s we could not construct PGI_Cog t_o apply in UK Biobank since we did not have an independent GWAS for cognitive ability in adults that excluded this cohort. The square brackets indicate significant comparisons highlighted in the text (z tests).

**Extended Data Figure 8.**
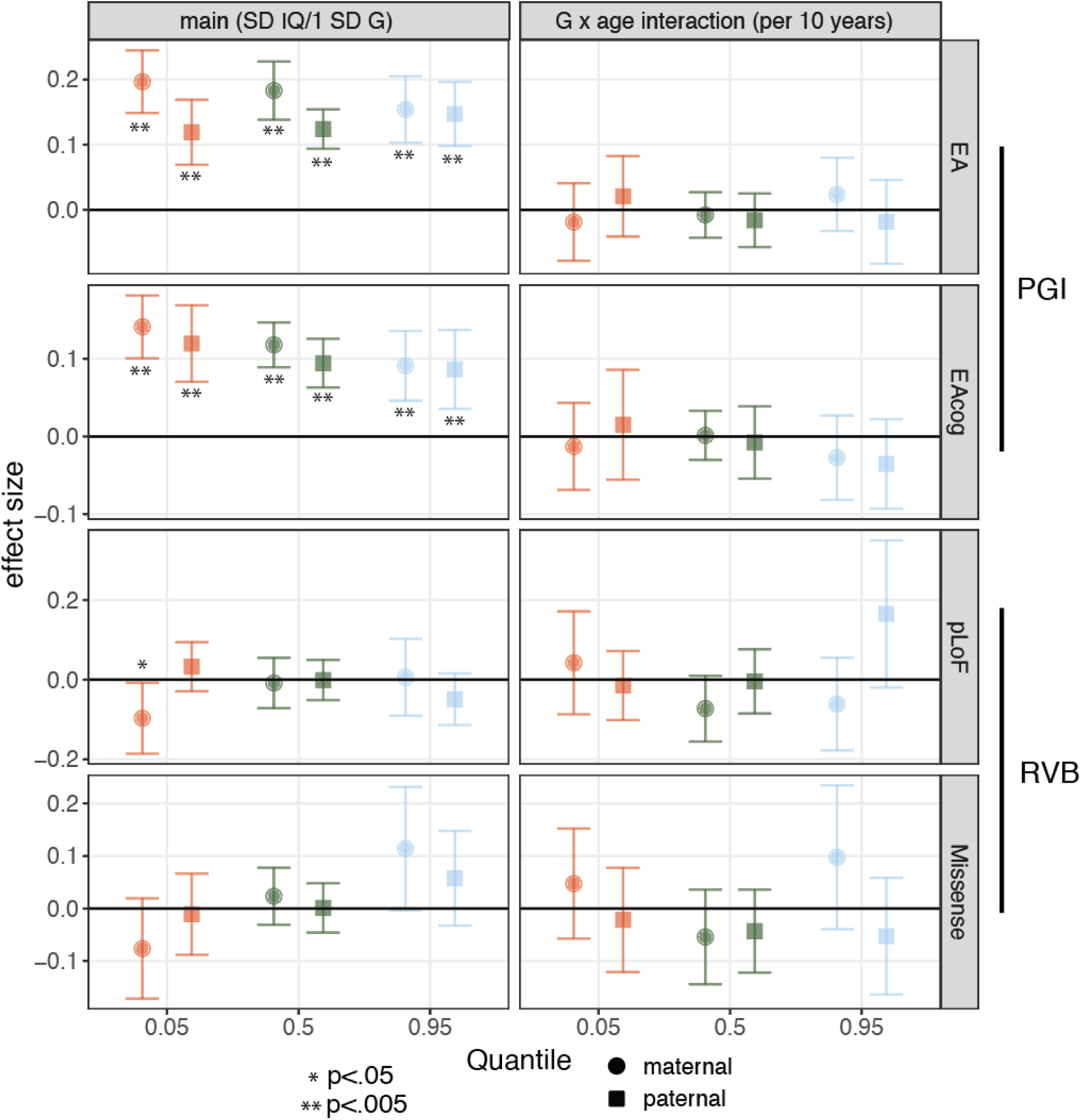
Influence of parental common and rare variants on different quantiles of the IQ distribution post-imputation. Standardized effects and 95% confidence intervals for quantile regression of the 5th, 50th, and 95th percentiles estimated from mixed-effects modeling with post-imputation IQ at ages 4, 8, and 16 for parental PGI_EA,_ PGI_Cog,_ RVB_pLoF a_nd RVB_Missense e_stimated in a trio model.

**Extended Data Figure 9.**
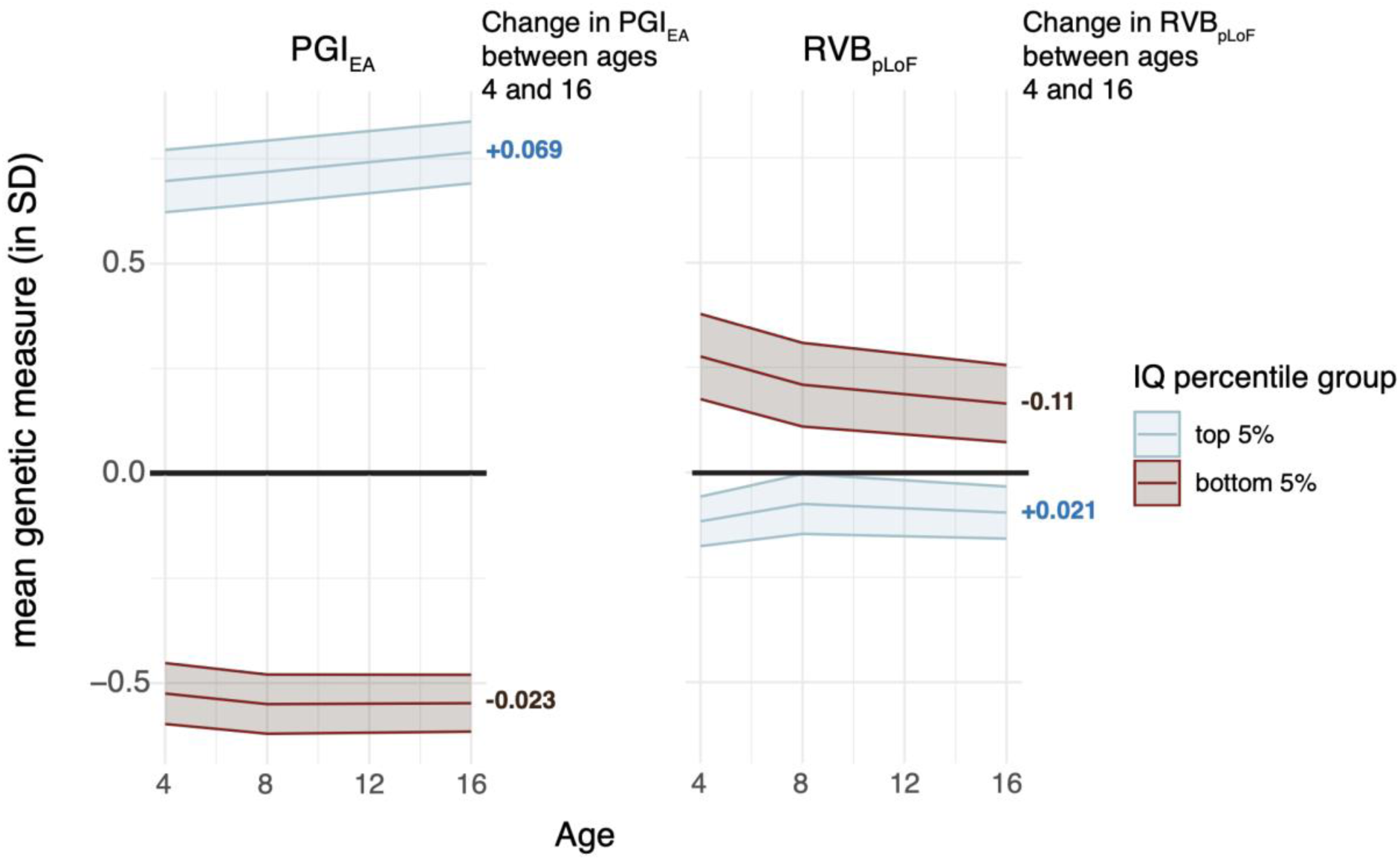
Summary of genetic trajectories across development, as inferred in this work. Mean PGI_EA (_left) and RVB_pLoF (_right) for individuals in the top (light blue) or bottom (brown) 5% percentile of IQ at a given age (inferred post-imputation of IQ). Note that the individuals in the top and bottom 5th percentile groups vary across development. Children at the top 5th percentile of IQ at age 16 have a higher mean PGI_EA t_han those at the top of the distribution at age 4, while those at the bottom 5th percentile have a relatively steady mean PGI_EA._ In contrast, the quantile-specific age interactions for RVB_pLoF s_uggest that children in the bottom 5th percentile of IQ at age 4 have a much higher burden of damaging rare variants than those at age 16, while those at the top 5th percentile have a stable and low rare variant burden.

**Extended Data Figure 10.**
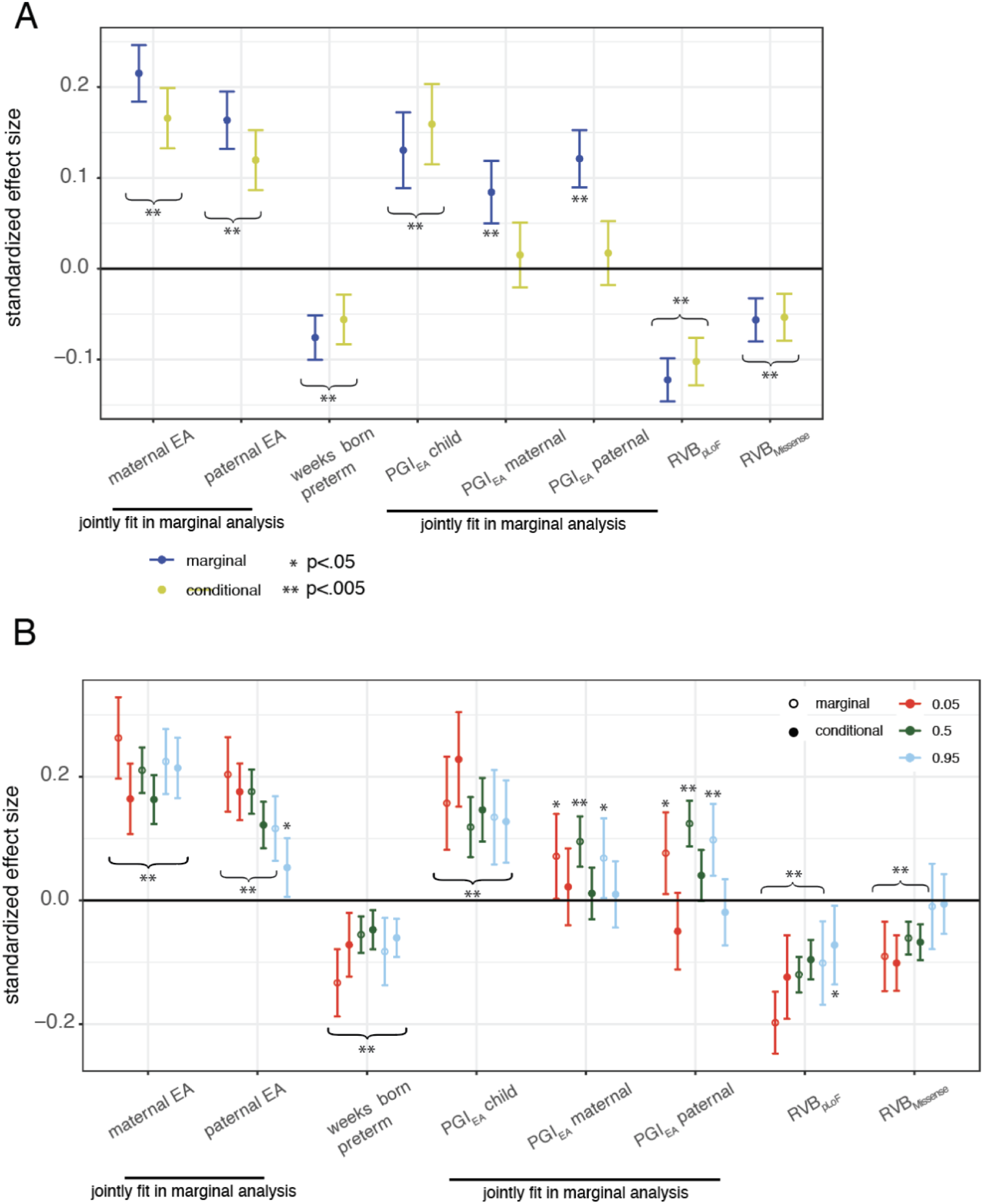
Associations between genetic and other factors and cognitive performance across ages in MCS. A) Standardized effects and 95% confidence intervals from obtained from two linear models, as indicated in the key: marginal associations from models in which only the indicated variable(s) were included in the model (in addition to standard covariates), and conditional associations from models in which all the variables were included in the same model. B) As in (A) using a quantile regression model for the quantiles as indicated in the figure. Asterisks indicate whether the estimate is significantly different from 0; note that curly brackets with asterisks indicate that the estimates spanned by the bracket are all significant.

