## Supplementary material for "The differential effects of common and rare genetic variants on cognitive performance across development"

### Supplementary Methods

#### *De novo* variant calling in ALSPAC and MCS

We carried out DNM calling in 1,326 trios in ALSPAC and 3,106 trios in MCS. Starting with the unfiltered callset obtained by GATK (described in detail in our recent data note[1]), a set of candidate *de novo* mutations (DNMs) was generated using `bcftools +trio-dnm2 --use-NAIVE` (http://samtools.github.io/bcftools/trio-dnm.pdf). In this mode, the program detects variants that violate the Mendelian patterns of inheritance simply by comparing parental and proband genotypes. However, a callset produced this way is dominated by false positives due to sequencing, mapping or alignment artifacts. In order to filter such artifacts, the following pipeline was used (Figure S7A):

1. Per-trio genotyping. We generated the following annotations at each candidate site using `bcftools mpileup -pa AD,QS,SP,SCR,FMT/NMBZ`:
    - AD, allelic depth, the number of reference and alternate reads observed in the three samples
    - QS, phred-score allele quality sum, an auxiliary annotation used by `bcftools +trio-dnm2`
    - SP, phred-scaled strand bias P-value
    - SCR, number of soft-clipped reads
    - NMBZ, Mann-Whitney U-z test of number of mismatches within supporting reads
2. Parental genotyping. We generated profiles of parental variant allele fraction (VAF) to identify problematic regions, as described in the section on “Parental VAF Profiles” below.
3. Obtaining posterior probabilities. We ran `bcftools +trio-dnm2 --strictly-novel --ppl` and `bcftools +trio-dnm2 --strictly-novel` to obtain posterior *de novo* probabilities using the DeNovoGear model[2] implemented in the trio-dnm2 plugin (http://samtools.github.io/bcftools/trio-dnm.pdf).
4. Random forest. We used random forest to score the candidate variants based on the following covariates:
    - DNM, the trio-dnm2 score
    - DNG, the DeNovoGear score
    - VCF_QUAL, variant quality as presented in the VCF QUAL column
    - MaxParentalVAF, maximum variant allele frequency (VAF) observed across parents at this site
    - NMBZ, Mann-Whitney U-z test of number of mismatches in variant reads
    - SCR, number of soft-clipped reads
    - SCBZ, Mann-Whitney U-z test of number of soft-clips in variant reads
    - MQBZ, Mann-Whitney U-z test of mapping quality of variant reads

    The random forest classifier was trained on a subset of calls manually inspected in IGV (Integrative Genomics Viewer)[3], separately for SNVs, deletions and duplications, and for autosomal and sex chromosomes in male and female samples. It was run with 100 trees in the forest and the relative contributions of the classification annotations are shown in Figure S7B.

5. Iterative manual curation and retraining of the random forest. We randomly selected 100 variants and curated them manually to extend the truth set, then re-trained the random forest classifier. We repeated this and the previous step 3-4 times until the random forest quality score separates clearly real from clearly false positive DNMs well.
6. We applied the following final filters:
   - In both ALSPAC and MCS
     - exclude any variants with population allele frequency 1% or higher (gnomAD v2.1)
     - exclude variants with parental depth lower than 8 reads
     - filter variants observed across unrelated parents, assessed with the LNP_VAF metric (log-normal probability of matching parental VAF profiles; i.e. we required LNP_VAF > -1000, see the section on “Parental VAF Profiles” below)
   - In ALSPAC
     - require random forest score bigger than 0.92
     - restrict C>A and G>T calls that passed stringent QC applied to the whole-exome data, due to a known sequencing artifact in the dataset[1]
   - In MCS
     - require random forest score bigger than 0.6
     - require VAF >= 25%

Note that different filters were applied in the two cohorts since they were differentially affected by a sequencing artifact described in the data note[1].

#### Parental VAF Profiles

The LNP_VAF annotation (log-normal probability of matching parental variant allele fraction profiles) is intended to filter sites in difficult regions by comparing the VAF profile in unrelated parents with the expected distribution. It is assumed that alternate reads in parents unrelated to the index proband represent sequencing errors. The method identifies a set of high-confidence *de novo* variants (i.e. sites with DNM score equal to 0) with sufficient coverage in all three trio samples (>=15x), proband VAF > 30%, and no alternate reads in the index proband's parents. For each such site, a VAF distribution is collected across all parents in the dataset who are unrelated to the proband of interest, and the mean $\mu$ and variance $\sigma$ are determined across all sites for each bin of the distribution. The log-normal score is then calculated as follows:
LNP_VAF = $-\sum_{i=1}^{N}(x_i - \mu_i)^2/\sigma_i^2$ , where N=50 is the number of bins of the VAF distribution.
It was then used in the filtering described above.

#### DNM callset summary and evaluation

The expected numbers of DNMs in different consequence classes were estimated per-gene for MANE transcripts from a sequence context-based mutational model [4]. The cumulative rates of expected DNMs per parental genome were as follows:

- synonymous: 0.160734,
- missense: 0.364192,
- loss-of-function, including stop gained, stop lost, start lost, splice acceptor, and splice donor consequence predictions: 0.0277504,
- stop gain: 0.0181173,

- frameshift:1.2*expected number of stop gain

In calculating the total expected number of DNMs in the dataset, the proportion of male vs female samples was taken into account as follows: 2*nSamples*dnm_rate for autosomal chromosomes and (nMales+2*nFemales)*dnm_rate for chrX. Pseudoautosomal regions were not considered in the calculation.

The mutation spectrum and distribution of parental VAF per dataset are shown in Figure S8, and the comparison of observed to expected DNMs in each consequence class in Figure S9. The number of DNMs in any one class or dataset did not differ significantly from the expected number, with the exception of missense mutations of which we observed a slight excess in MCS.

#### Gene set associations and enrichment

To derive gene set-specific associations (Extended Data Figure 5, Figure S5, Figure S6), we conducted mixed-effect linear modeling identically as above, restricting the $RVB_{pLoF}$ calculation to genes only present in a given gene set. To make the effects comparable between gene sets of varying sizes, we divided the effects by the number of genes in the gene set.

To derive gene set-specific enrichment for a given gene set *g* of size *n*, we randomly sampled *n* genes not in *g* to generate a matched gene set *h*. To ensure that the distribution of heterozygous PTV selection coefficients in *h* is similar to *g*'s, we first calculated the 5-quantiles of the selection coefficient distribution of *g* and sampled *n*/5 genes from each quantile, rounded to the nearest upper whole integer. We then calculated a *h* specific $RVB_{pLoF}$ and repeated the analyses above, repeating this procedure 100 times. We then combined the estimates of the 100 gene sets using Rubin's rule. Enrichment was calculated as the ratio of the $RVB_{pLoF}$ effects for *g* and the pooled effects for *h*. Significance was determined using a Wald test.

### Supplementary Tables

Table S1

| Factor loading | Key stage 2 (ALSPAC) | Key Stage 3 (ALSPAC) |
|---|---|---|
| Math | 0.87 | 0.91 |
| Science | 0.89 | 0.94 |
| English | 0.81 | 0.77 |
| Proportion variance explained | 0.74 | 0.77 |

Key stage factor loadings and proportion of variance explained by one factor model on academic performance in ALSPAC.

Table S2

| IQ measure | Heritability | $r_g$ with age 4 | $r_g$ with age 8 | n | IQ imputation status |
|---|---|---|---|---|---|
| Age 8 | 0.46 (0.154) | - | - | 3,176 | pre-imputation |
| Age 16 | 0.73 (0.153) | - | 0.96 (0.042) | 3,176 | pre-imputation |
| Age 4 | 0.46 (0.081) | - | - | 6,495 | post-imputation |
| Age 8 | 0.48 (0.080) | 0.97 (0.021) | - | 6,495 | post-imputation |
| Age 16 | 0.56 (0.080) | 0.96 (0.024) | 0.96 (0.031) | 6,495 | post-imputation |

Heritabilities and genetic correlations of the IQ measures in ALSPAC pre- and post-imputation, estimated using GREML-LDMS[5], with standard errors shown in brackets. Genetic correlations were estimated between post-imputation IQ measures or between pre-imputation IQ measures. GREML-LDMS estimates of heritability at age 4 pre-imputation are missing from this table as they did not converge. Estimates of heritabilities and genetic correlation between IQ at age 8 and 16 pre-imputation were calculated in unrelated individuals that had both measures.

Table S3

| IQ measure | $r_g$ with EA | $r_g$ with Cog | $r_g$ with NonCog |
|---|---|---|---|
| Age 4 | 0.903 (0.065) | 0.863 (0.062) | 0.416 (0.077) |
| Age 8 | 1.118 (0.081) | 0.952 (0.071) | 0.629 (0.089) |
| Age 16 | 0.977 (0.068) | 0.878 (0.061) | 0.502 (0.076) |

Genetic correlations of the post-imputation IQ measures in ALSPAC with external GWASs using LDSC[6], with standard errors in brackets.

Table S4

PGI effects for cross-sectional and mixed-effects models on IQ in ALSPAC. First column indicates the PGI being assessed in the regression, second column indicates whether the effect is the proband's population or direct effect estimate or the paternal/maternal effect estimate from a trio model, columns three through five indicate the effect size, standard error, and the p value, respectively, the sixth column indicates whether it was a cross-sectional or mixed-effects model, the seventh column indicates for a cross-sectional model the age at which the IQ test was administered or for a mixed-effects model whether it is an estimate of the main or age interaction effect, the eighth column indicates whether the regression was conducted before or after imputation of IQ measures, and the ninth column indicates whether the regression was conducted in the full cohort or the trio subset.

Table S5

PGIs effects on academic performance in ALSPAC. First column indicates the PGI being assessed in the regression, second column indicates whether the effect is the proband's population or direct effect estimate or the paternal/maternal effect estimate from a trio model, columns three through five indicate the effect size, standard error, and the p value, respectively, the sixth column indicates the year at which the exams were administered or whether it is the age interaction estimated by taking the difference the scaled academic performance measures.

Table S6

RVB effects for cross-sectional and mixed-effects models on IQ in ALSPAC. First column indicates the RVB being assessed in the regression, second column indicates whether the effect is the proband's population or direct effect estimate or the paternal/maternal effect estimate from a trio model, columns three through five indicate the effect size, standard error, and the p value, respectively, the sixth column indicates whether it was a cross-sectional or mixed-effects model, the seventh column indicates for a cross-sectional model the age at which the IQ test was administered or for a mixed-effects model whether it is an estimate of the main or age interaction effect, the eighth column indicates whether the regression was conducted before or after imputation of IQ measures, and the ninth column indicates whether the regression was conducted in the full cohort or the trio subset.

Table S7

RVB effects on academic performance in ALSPAC. First column indicates the RVB being assessed in the regression, second column indicates whether the effect is the proband's population or direct effect, columns three through five indicate the effect size, standard error, and the p value, respectively, the sixth column indicates the year at which the exams were administered or whether it is the age interaction estimated by taking the difference the scaled academic performance measures.

Table S8

Quantile regression cross-sectional and mixed-effects model results for PGI and RVB effects on IQ in ALSPAC. First column indicates the PGI (EA, Cog, or Noncog) /RVB (pLoF or Missense) being assessed in the regression, second column indicates whether the effect is the proband's population or direct effect estimate or the paternal/maternal effect estimate from a trio model, columns three through five indicate the effect size, standard error, and the p value, respectively, the sixth column indicates whether it was a cross-sectional or mixed-effects model, the seventh column indicates for a cross-sectional model the age at which the IQ test was administered or for a mixed-effects model whether it is an estimate of the main or age interaction effect, the eighth column indicates the quantile being assessed, the ninth column indicates whether the regression was conducted before or after imputation of IQ measures, and the tenth column indicates whether the regression was conducted in the full cohort or the trio subset.

Table S9

Quantile regression and difference in effects for PGI and RVB effects on academic performance in ALSPAC. First column indicates the PGI (EA, Cog, or Noncog) /RVB (pLoF or Missense) being assessed in the regression, second column indicates whether the effect is the proband's population or direct effect estimate or the paternal/maternal effect estimate from a trio model, columns three through five indicate the effect size, standard error, and the p value, respectively, the sixth column indicates the year at which the exams were administered or whether it is the age interaction estimated by taking the difference the scaled academic performance measures, the seventh column indicates the quantile being assessed.

### Supplementary Figures

Figure S1

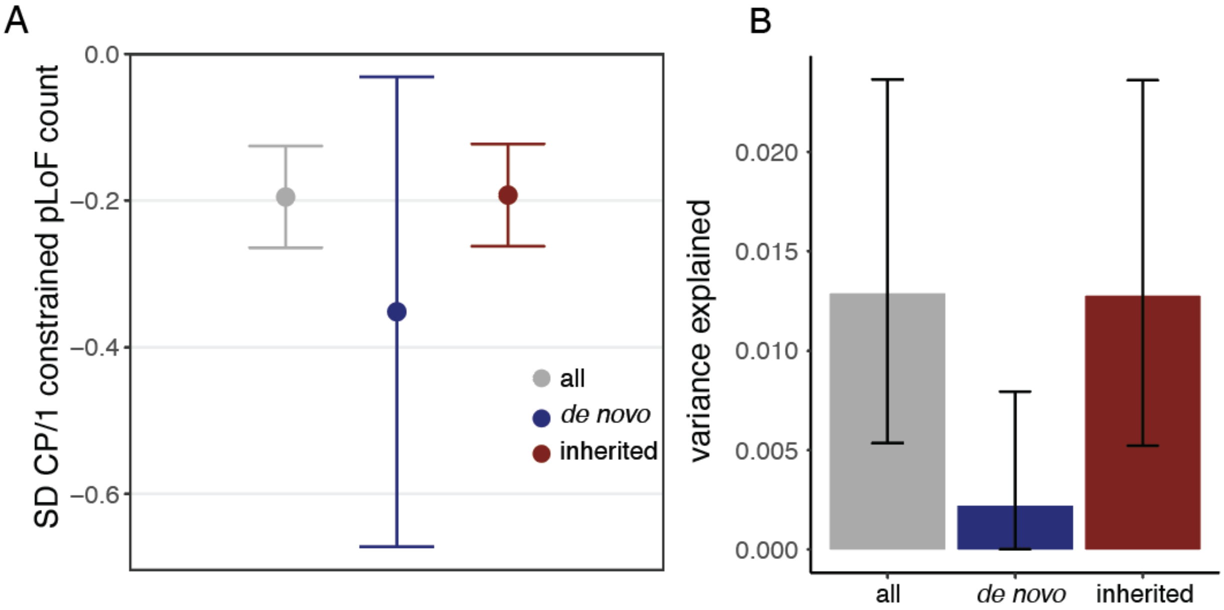


Association between inherited and *de novo* pLoF variant counts in constrained genes and composite cognitive performance scores in MCS. A) Standardized effects for the main and RVB-by-age interaction effect from a longitudinal mixed-effects model of constrained pLoF counts on standardized cognitive performance scores when considering all variants or *de novo* mutations and inherited variants separately. B) Variance explained by constrained pLoF counts when considering all variants or *de novo* and inherited separately.

Figure S2

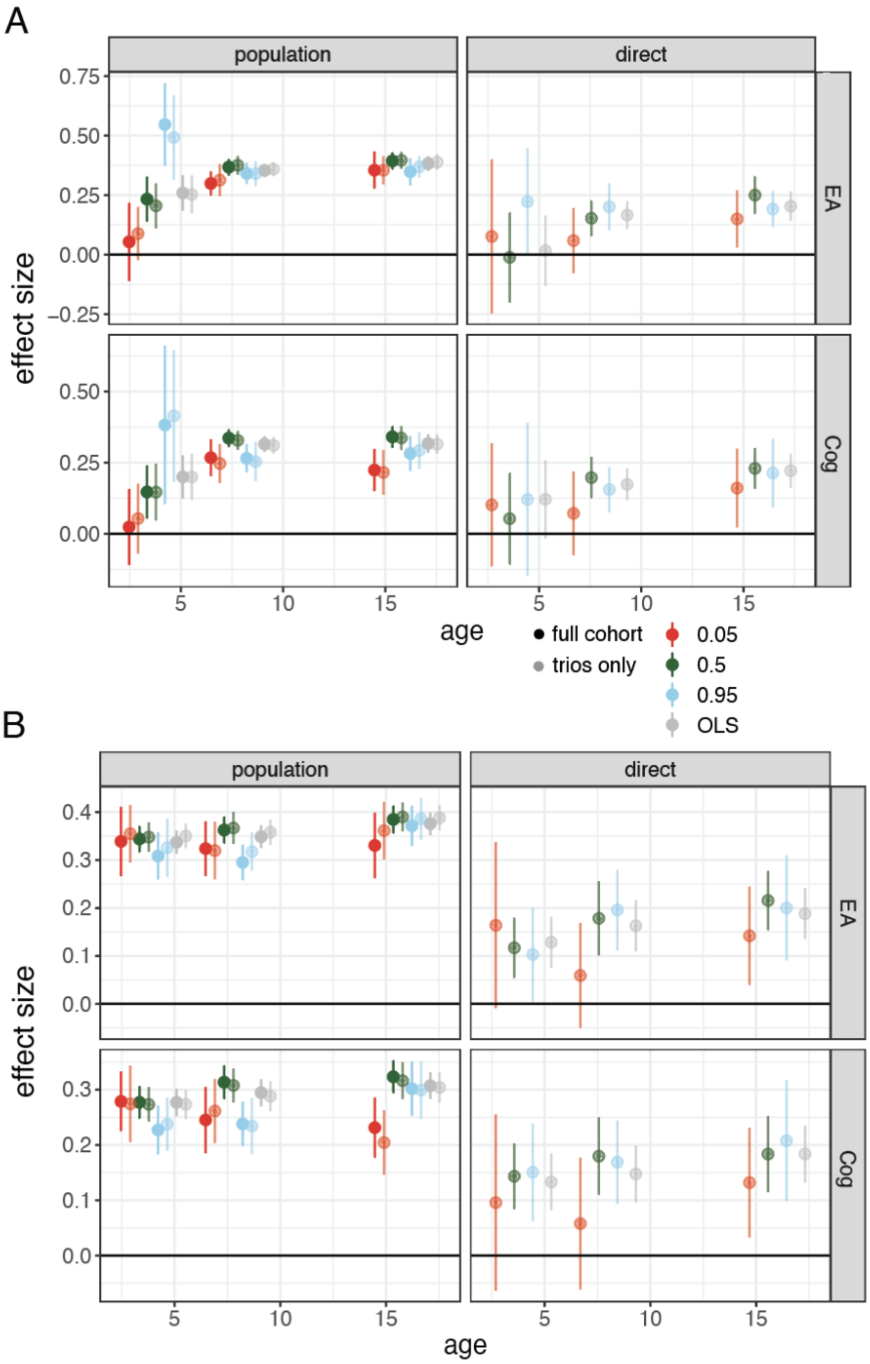


Influence of PGIs on different quantiles of the IQ distribution in ALSPAC estimated by applying quantile regression in cross-sectional analyses. A) Results using pre-imputation IQ measures. Standardized effects and 95% confidence intervals for quantile regression of the 5th (red), 50th (green), and 95th (blue) percentiles and ordinary least squares (OLS) linear regression (gray) estimated from cross-sectional associations with pre-imputation IQ at ages 4, 8, and 16 for $PGI_{EA}$ and $PGI_{Cog}$ before (left) and after (right) controlling for parental genetic measures. B) Same as (A) using post-imputation IQ measures.

Figure S3

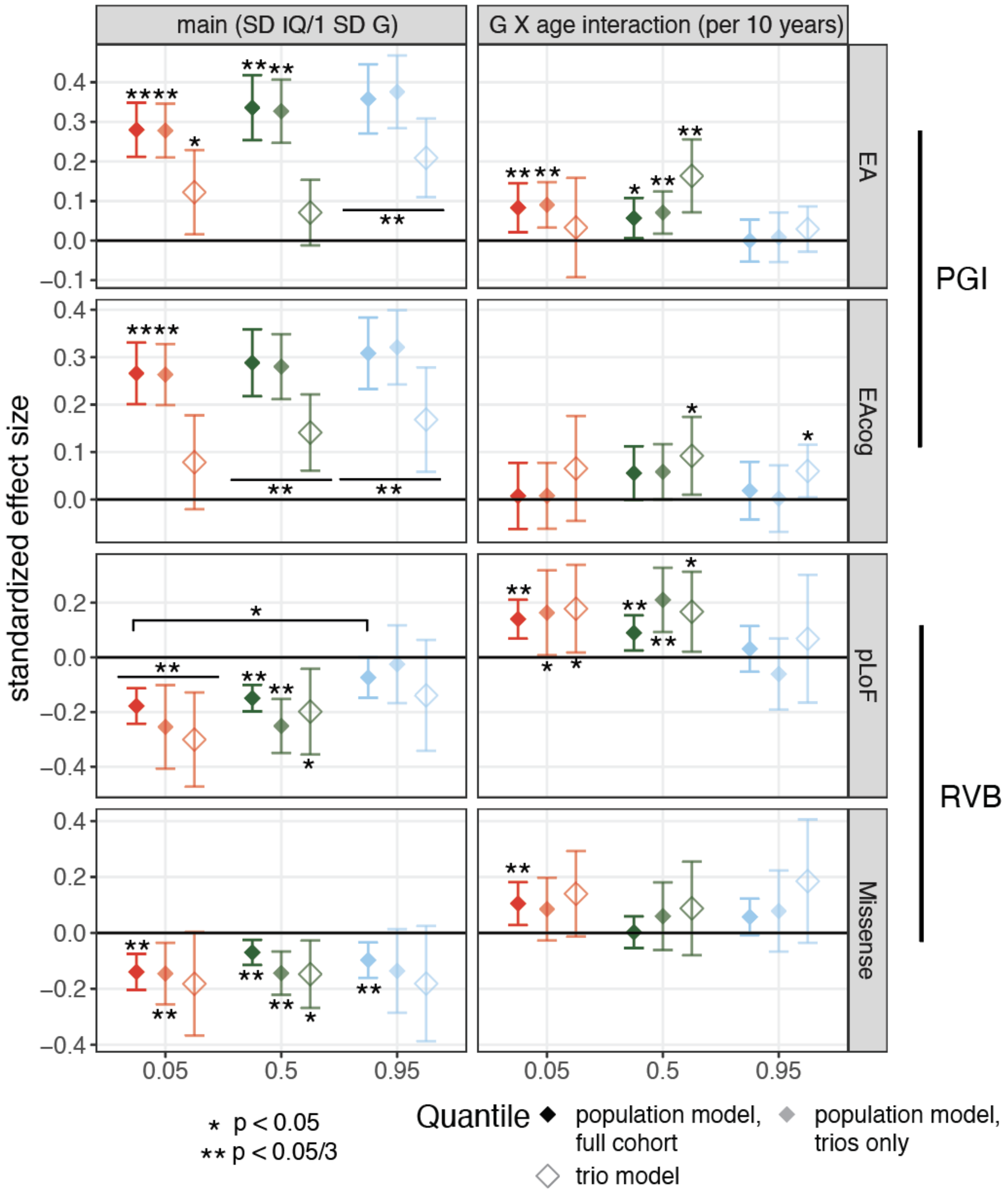


Influence of common and rare variants on different quantiles of the IQ distribution pre-imputation. (As for Figure 3BC, which is based on post-imputation IQ.) Standardized effects and 95% confidence intervals for quantile regression of the 5th, 50th, and 95th percentiles estimated from mixed-effects modeling with pre-imputation IQ at ages 4, 8, and 16 for $PGI_{EA}$, $PGI_{Cog}$, $RVBS_{pLoF}$ and $RVBS_{Missense}$ before (left) and after (right) controlling for parental genetic measures. The square brackets indicate significant comparisons highlighted in the text (z tests).

Figure S4

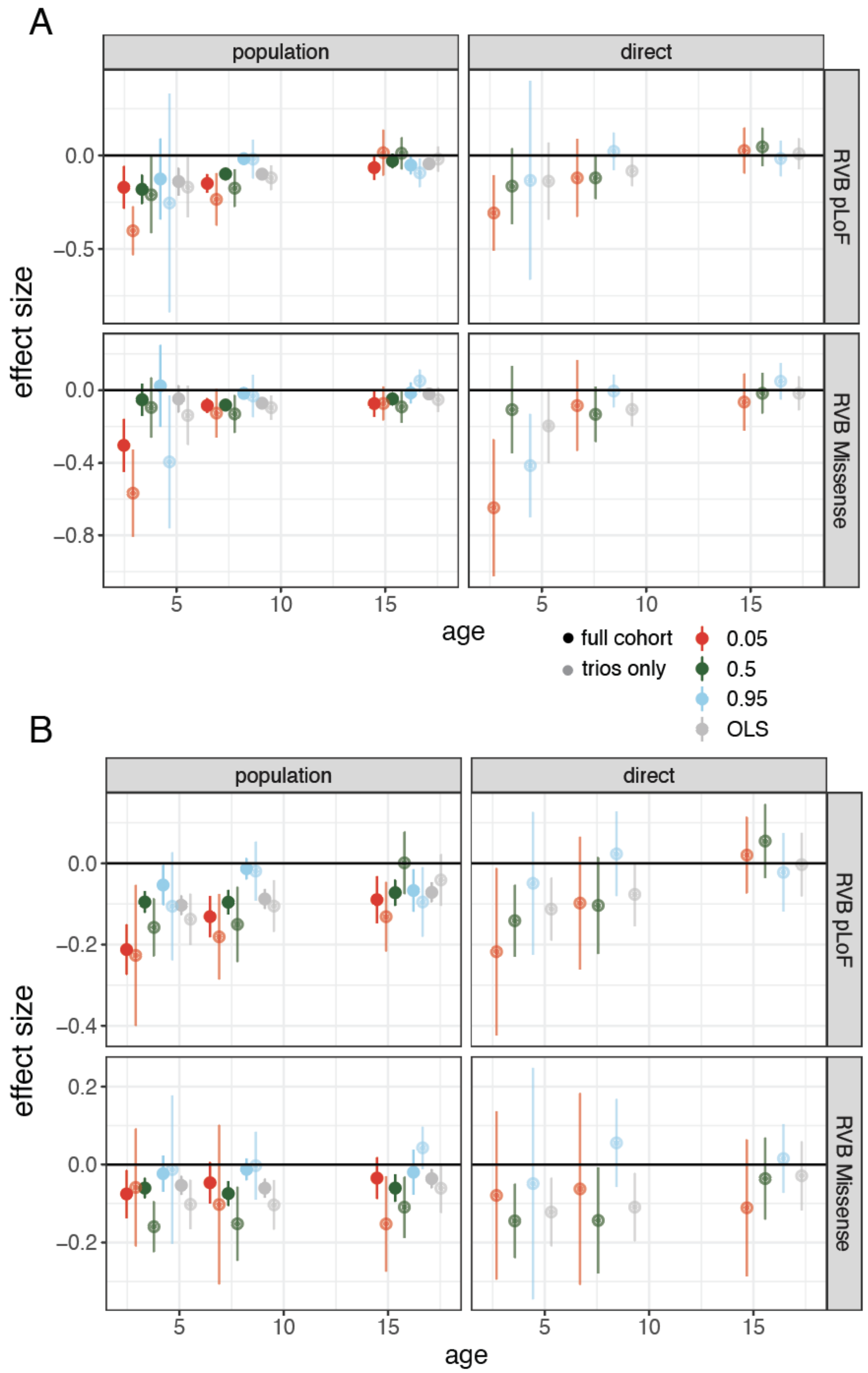


Influence of RVB on different quantiles of the IQ distribution using pre-imputation IQ measures in ALSPAC, from cross-sectional analyses. A) Standardized effects and 95% confidence intervals for quantile regression of the 5th (red), 50th (green), and 95th (blue) percentiles and ordinary least squares (OLS)

regression (gray) estimated from cross-sectional associations with pre-imputation IQ at ages 4, 8, and 16 for $PGI_{EA}$ and $PGI_{Cog}$ before (left) and after (right) controlling for parental genetic measures. B) Same as (A) using post-imputation IQ measures.

Figure S5

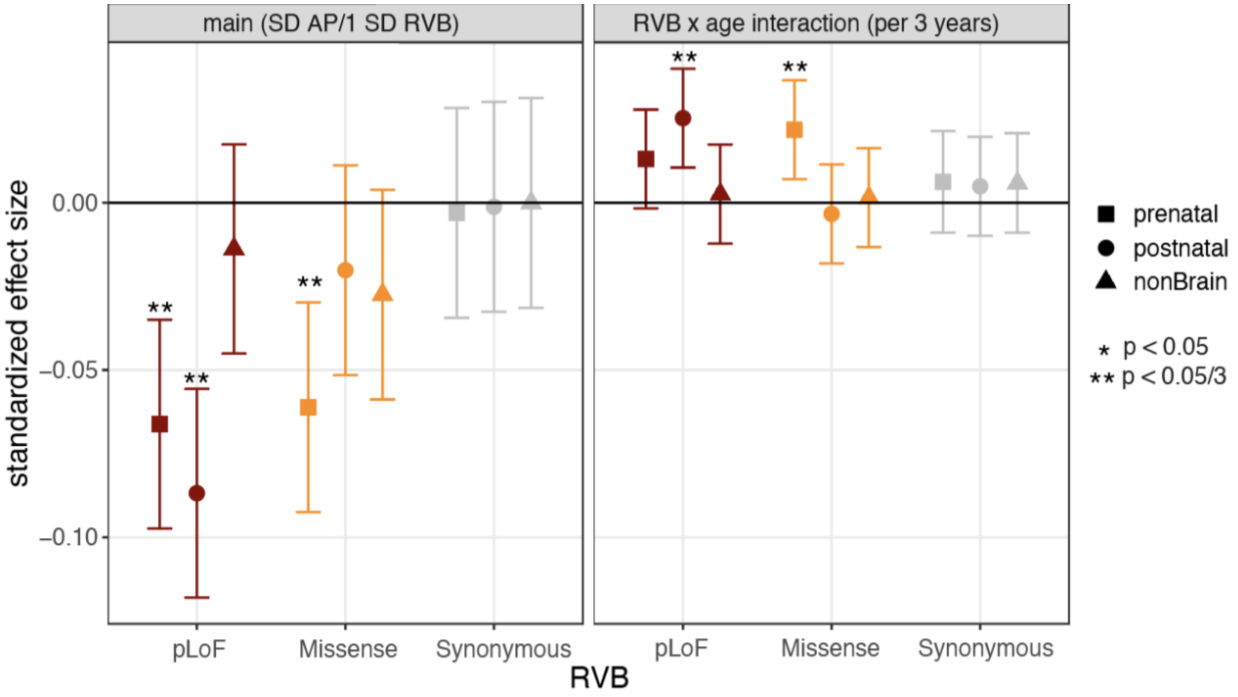


Associations between RVB in gene sets defined according to their expression patterns and academic performance in ALSPAC. (Similar to Extended Data Figure 5A which shows results for IQ in ALSPAC.) Standardized effects and 95% confidence intervals estimated for main effects on academic performance and RVB-by-age-interaction effects for each RVB in three mutually exclusive gene sets from Li et al. These comprise genes in co-expressed clusters that are more highly expressed in prenatal or postnatal brain or that are not detected in the study (non-brain).

Figure S6

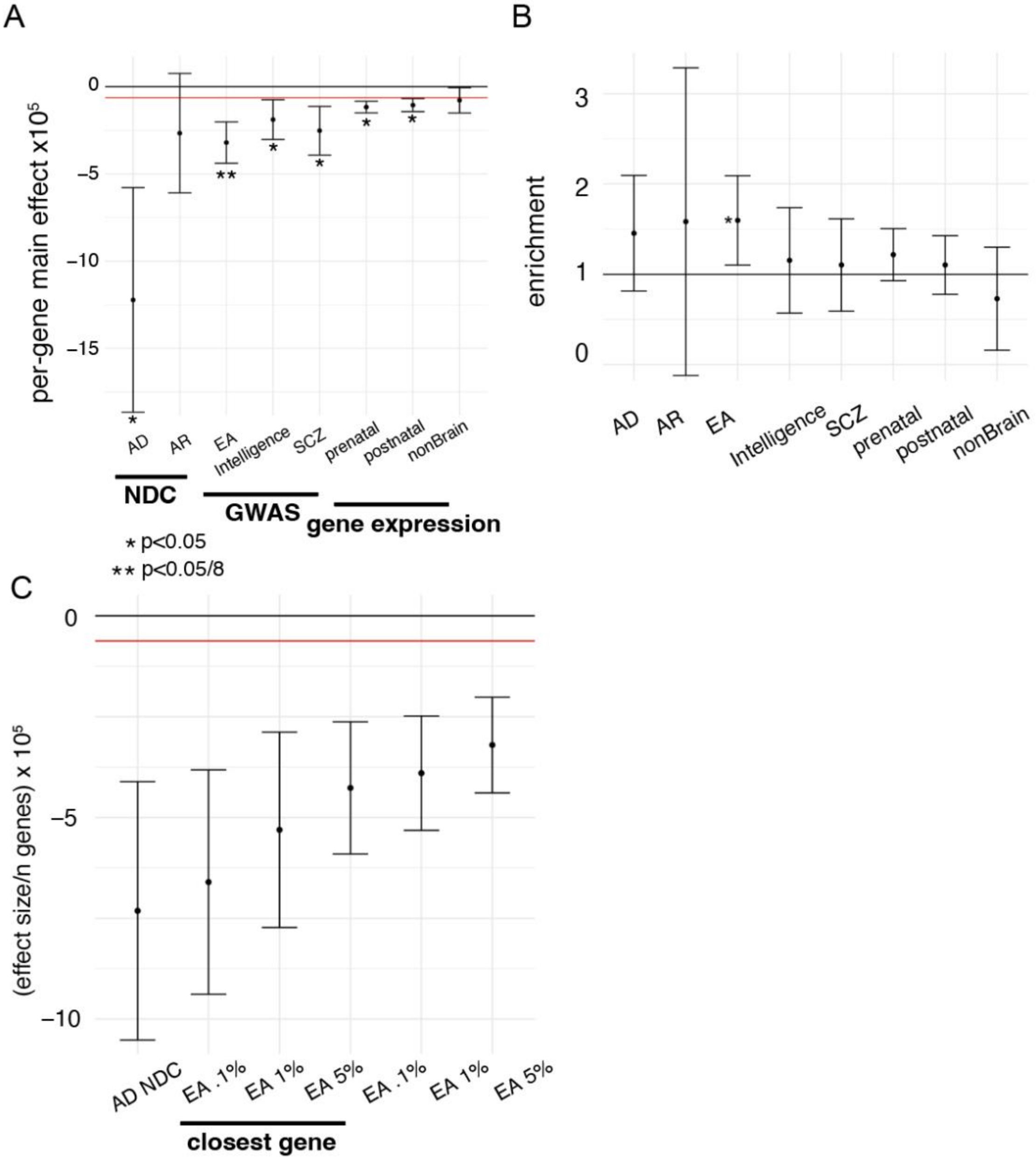


Replication of effects of gene set-specific RVB on composite cognitive performance measure in MCS. (Similar to Extended Data Figure 5BDE which show results using the main effect from mixed-effect models fitted on IQ in ALSPAC.) A) Effects of $RVB_{pLoF}$ on composite cognitive performance score stratified by gene set, divided by the number of genes in the given gene set, with 95% confidence intervals. Red horizontal line indicates the average effect for $RVB_{pLoF}$ across all genes. Asterisks indicate the p-value for difference in per-gene effects between a given gene set and all genes using a z test, with * indicating nominal significance and ** indicating Bonferroni significance for 8 tests. B) Ratio of the main effect for $RVB_{pLoF}$ for the indicated gene set relative to randomly sampled gene sets with matching underlying $s_{het}$ distributions (enrichment). C) Per-gene main effect for $RVB_{pLoF}$ using different gene FDR threshold cutoffs based on gene prioritization in Lee et al. and by restricting to prioritized genes at a given cutoff that are also the nearest genes to the prioritizing SNP in the Lee et al. GWAS[7]. AD/AR NDC: Autosomal dominant/recessive neurodevelopmental disorder genes with loss-of-function mechanism from DDG2P[8], EA 5%: educational attainment GWAS prioritized genes by Lee et al.[7] at 5% FDR threshold, IQ: intelligence GWAS prioritized genes from Savage et al.[9], SCZ: schizophrenia GWAS prioritized genes from Pardiñas et al.[10]

Figure S7

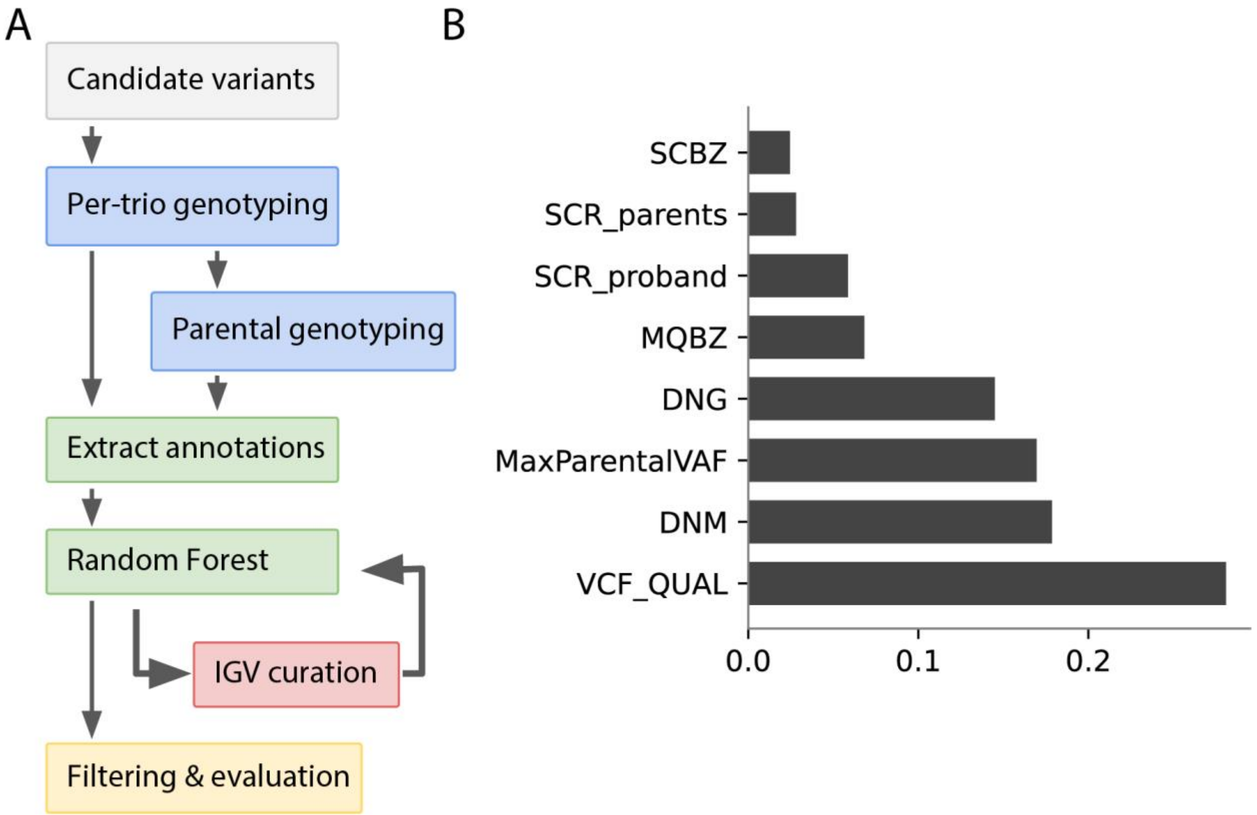


Calling and quality control of *de novo* mutations. A) Flow diagram illustrating the calling and quality control process. B) Relative importance of features included in the random forest model used for quality control. SCBZ: Mann-Whitney U-z test of number of soft-clips in variant reads; SCR: number of soft-clipped reads; MQBZ: Mann-Whitney U-z test of mapping quality of variant reads; DNG: DeNovoGear score; MaxParentalVAF: maximum variant allele frequency (VAF) observed across parents at this site; DNM: trio-dnm2 score; VCF_QUAL: variant quality as presented in the VCF QUAL column from GATK.

Figure S8

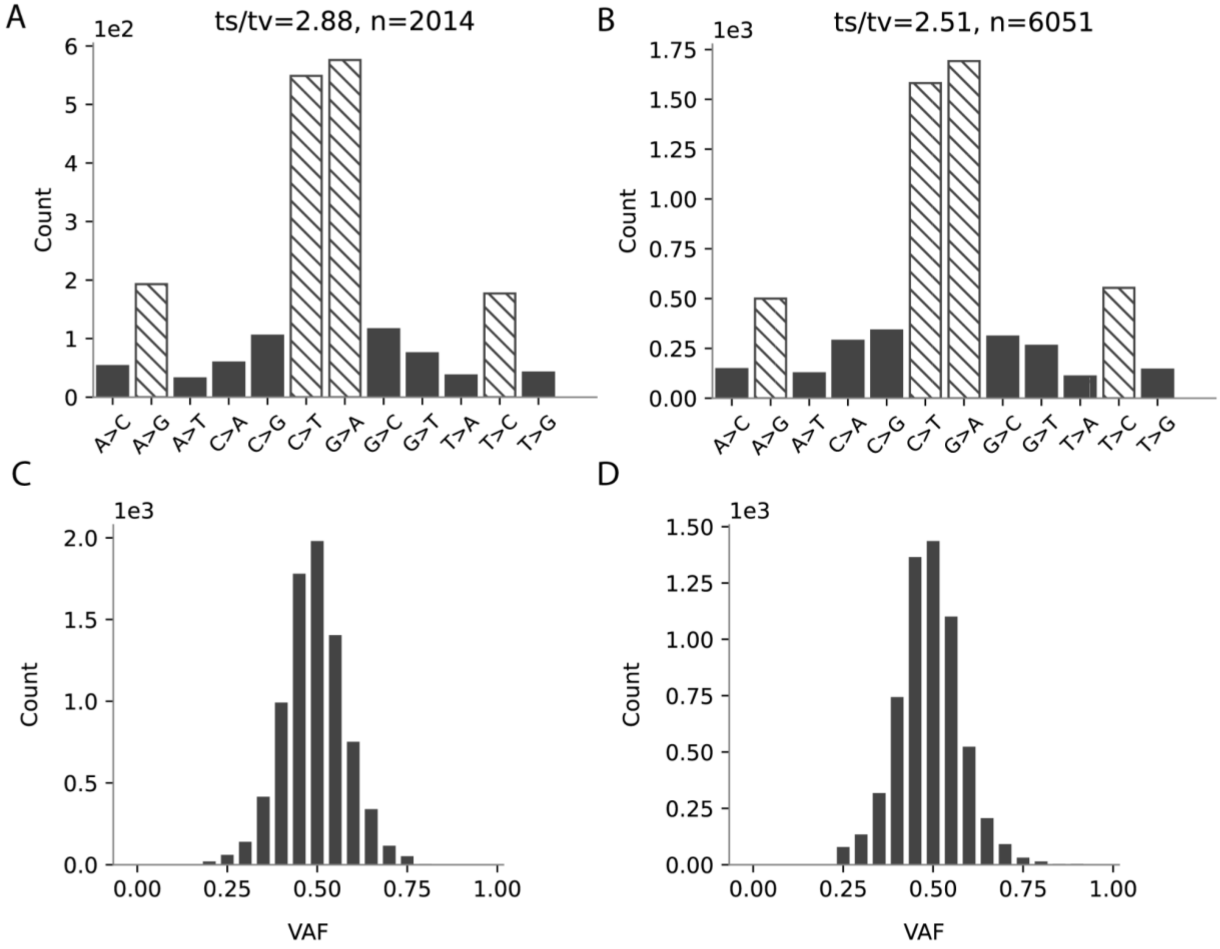


*De novo* mutation spectrum for single nucleotide variants in ALSPAC (A) and MCS (B). The hashed bars are transitions and the black bars transversions. ts/tv = transition/transversion ratio. Distribution of variant allele fraction (VAF) in ALSPAC (C) and MCS (D).

Figure S9

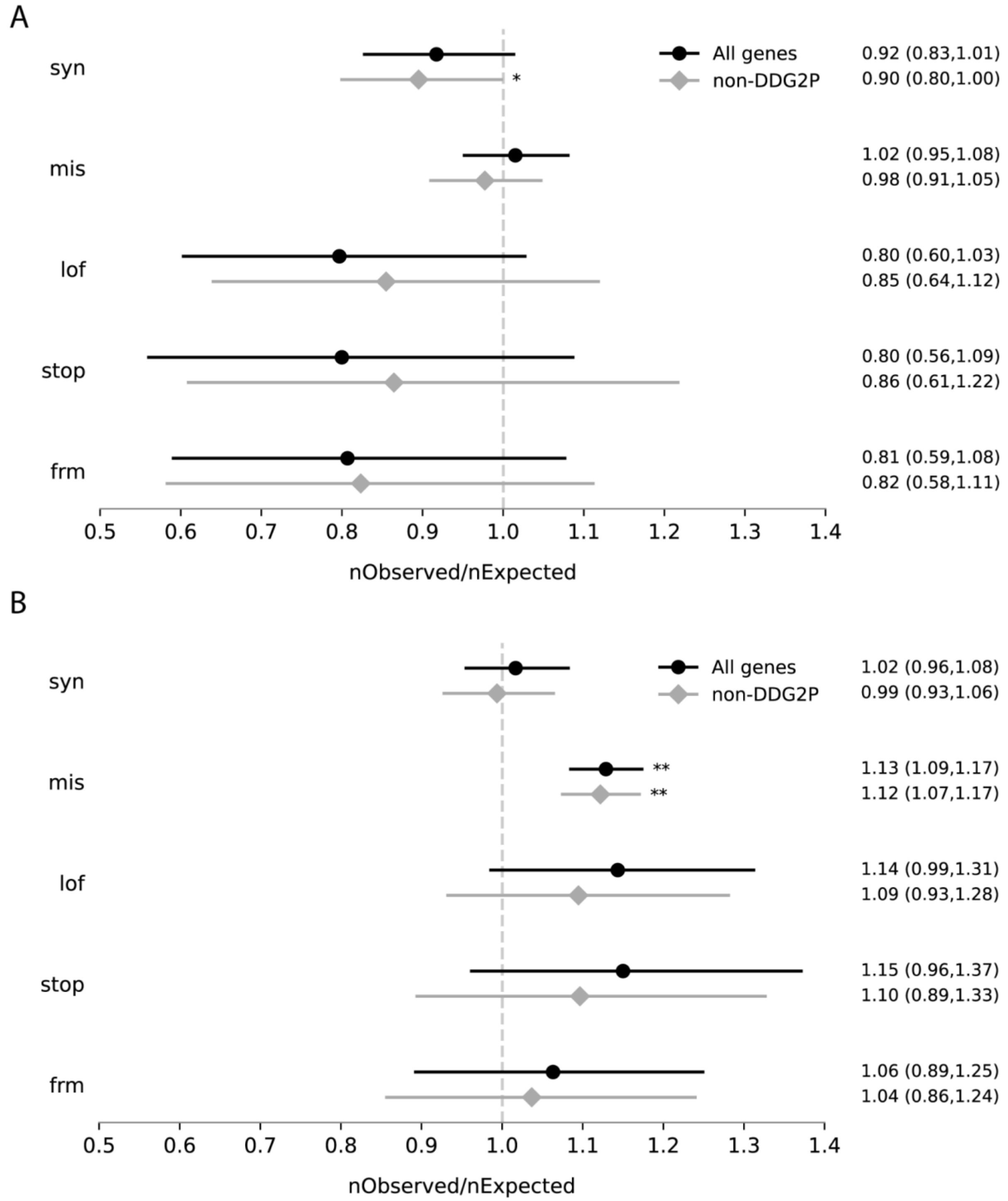


Observed vs expected number of de novo mutations in ALSPAC (A) and MCS (B) with 95% confidence intervals and Poisson test significance threshold of $p<0.05$ (*) and $p<0.01$ (**). The considered mutations types were: synonymous (syn), missense (mis), loss of function (lof), stop gained (stop) and frameshift indels (frm).

### Supplementary Note 1: IQ imputation in ALSPAC

#### Imputing IQ using SoftImpute

IQ was measured in ALSPAC at ages 4, 8, and 16 via psychometric testing administered in an in-person visit. Of the 8,804 children in the study with SNP array data, 6,496 had IQ measured at least once, with the lowest attendance at age 4 and relatively higher attendance at ages 8 and 16 (Extended Data Figure 1A). IQ measures were highly correlated between ages, with pairwise correlations ranging from 0.51 (between ages 4 and 16) to 0.63 (between ages 4 and 8).

In order to address nonresponse bias and increase our phenotyped sample size, we imputed missing IQ values across ages for all individuals with at least one IQ measurement and at least 20% of variables considered for the imputation having nonmissing values per individual using SoftImpute[11], an imputation algorithm that leverages the correlation structure across selected related variables to simultaneously impute all missing phenotype values. We first sought to identify a set of variables that maximize the imputation accuracy. We assessed three groups of variables: 1) the two other IQ measures, sex, parental income, and birth weight (base set), 2) the base set as well as a range of variables measured across life including additional cognitive and behavioral assessments (expanded set) (see Methods), and 3) the expanded set without the base set of variables (auxiliary set). To assess imputation quality for each of these three imputation strategies, we masked 100 measured IQ values at a given age and compared how the measured values compare to the imputed values.

The highest correlations were observed in the expanded set across ages (Extended Data Figure 1B). Importantly, we observed a median correlation of 0.61 for imputation accuracy for IQ at age 4, where we have the fewest phenotyped individuals. When using the auxiliary set of variables, the performance at age 4 and age 8 was similar to that when using the base set, but performance was markedly worse at age 16 with a median correlation difference of 0.24 ($p<.05$ Wilcoxon rank-sum test). These results suggest the auxiliary variables improved imputation most in the younger ages likely due to most of them being measured early in childhood, while at age 16 the base set contained the majority of the information that informed the imputation. Reassuringly, we observed consistent and accurate imputation across ages and used imputed IQ derived from the expanded set of variables throughout.

#### Common variant heritability and genetic correlations of IQ across time

We then compared the common variant genetic architecture of the imputed IQ values across ages. We conducted genome-wide association studies (GWASs) on IQ measurements at each age in an unrelated set of individuals inferred to have European genetic ancestry (n=6,495) (Methods). We used GREML-LDMS to calculate heritabilities and genetic correlations between the three GWASs. We found significant heritabilities for all traits, with $h^2$ estimates between 0.46-0.56 (Table S2). Additionally, we found significant genetic correlations between all of the traits, with none showing significant difference from an $r_g = 1$ (Table S2). These estimates of childhood IQ are concordant with previous estimates of SNP heritabilities ranging from 0.22-0.72[12,13].

We additionally used LD score regression to calculate genetic correlations between the three GWASs and external GWASs of EA[7] and the cognitive and non-cognitive components of EA (EA-Cog and EA-nonCog)[14]. We found all genetic correlations were significant, with the correlations for EA and EA-Cog being consistently high ($r_g > 0.85$) (Table S3). In contrast, the genetic correlations with EA-nonCog were lower (0.42-0.63) and significantly less than 1, consistent with the lower genetic correlation estimates between childhood IQ and EA-nonCog relative to that with EA-Cog previously observed[14]. Overall, these results indicated the common variant genetic architecture of measured+imputed IQ was highly similar across ages and to those of cognitive performance and educational attainment GWASs in directly-measured external samples.

#### Supplementary Note 2: Effects of polygenic scores and rare variant burden on academic performance measures in ALSPAC

As a complementary approach, we assessed the influence of the PGIs and RVBs on academic performance in ALSPAC. In Year 6 and Year 9 (roughly ages 11 and 14, respectively; known as Key Stages 2 and 3 in the UK) children were administered three standardized exams covering English, Mathematics, and Science from which we derived a composite academic performance metric which we showed was measuring the same latent construct across time (Table S1; Methods). For children who had complete data for the three exams at both ages (n=3,895), we assessed the contribution of the three PGIs to academic performance in a similar way to that described for IQ in the main text. We found significant ($p<1.1\times10^{-17}$) population effects for all PGIs and significant increases in effects with age for $PGI_{EA}$ and $PGI_{NonCog}$($p<2.1\times10^{-8}$) (Extended Data Figure 2, Table S5). In a trio analysis (n=3,024), we found evidence for direct genetic effects of $PGI_{EA}$ and $PGI_{Cog}$ on academic performance (Extended Data Figure 2), consistent with previous work in other cohorts[19]. Though all of the PGI-by-age interactions were positive, only the $PGI_{EA}$ showed a significant increase in direct effects (p=0.014), likely due to the reduction in power of the decreased sample size and the narrower age range considered. However, broadly these results for the impact of common genetic variation on academic performance mirror what we observed for IQ. For $RVB_{pLoF}$ and $RVB_{Missense}$, we found that they were significantly negatively associated with academic performance at age 11 but their effects attenuated with age (Extended Data Figure 4, Table S7).

#### Supplementary Note 3: Differential effects of rare variants in different gene sets on IQ

We assessed whether the effects of rare variants on IQ differed according to the expression patterns of the genes in which they lie. Prior GWAS studies suggest that genes overlapping loci associated with intelligence and educational attainment are enriched for expression in brain[7,9], as expected, and that genes associated with educational attainment showed on average relatively higher expression in prenatal rather than postnatal brain[7]. Similarly, genes implicated in autism by rare variants are also enriched for expression in prenatal brain [15], as are those associated with neurodevelopmental disorders[16]. We recalculated RVB stratified by genes' assignments to co-expressed modules enriched for expression in prenatal brain (n genes = 6,808), postnatal brain (6,078), or undetected in brain (3,028) [17]. We found that RVBs in the non-brain genes were not associated with IQ (Extended Data Figure 5A). In contrast, $RVB_{pLoF}$ and $RVB_{Missense}$ in the prenatal and postnatal gene sets were associated with IQ with main effects of similar magnitudes. In the prenatal gene set, $RVB_{pLoF}$ had a significant interaction with age pre- and post-imputation ($p<0.05/3$) and $RVB_{Missense}$ had a nominally significant interaction post-imputation ($p<0.05$). In contrast, we found only a nominally significant age interaction for $RVB_{pLoF}$ in postnatal genes post-imputation, and no evidence for an age interaction pre-imputation. Hence, rare damaging variants in genes preferentially expressed in the prenatal brain showed a clear attenuation of the rare variant effect with age on IQ, but there is only mixed evidence for an age effect among genes preferentially expressed in postnatal brain.

Similarly, we then assessed the association between RVBs stratified by expression patterns in the brain and academic performance in ALSPAC. As before, we detected no significant associations with any $RVB_{Synonymous}$ and or RVB calculated in genes not detected in brain (Figure S5). We found significant associations between $RVB_{pLoF}$ and $RVB_{Missense}$ calculated in prenatal genes and academic performance, and $RVB_{pLoF}$ calculated in postnatal genes. However, the patterns of attenuation were less clear: $RVB_{pLoF}$ only showed significant attenuation with age when calculated in postnatal genes and $RVB_{Missense}$ only showed a significant attenuation in prenatal genes. The confidence intervals on the age interaction estimates are large, particularly for the positive age interaction for $RVB_{pLoF}$ calculated in prenatal genes where the estimate was nearly nominally significant, suggesting statistical power is a limitation.

We next explored other gene sets that we hypothesized might have differential contributions to the $RVB_{pLoF}$ association with IQ. In addition to the aforementioned gene sets defined by expression pattern, we considered gene sets ascertained for severe monogenic autosomal dominant (n genes = 337) or recessive NDCs (636) and genes prioritized via common variant genome-wide association studies of EA[7] (1,838), cognitive ability (1,912), and schizophrenia[10] (1,558). We calculated $RVB_{pLoF}$ subsetted to only genes in a given gene set, and considered the main effect on IQ in ALSPAC estimated in a mixed-effects model, divided by the number of genes in the gene set, yielding per-gene effect estimates (Extended Data Figure 5B). Of the gene sets defined based on expression in the brain, only the prenatal gene set showed a modestly greater per-gene effect than the exome-wide average (p=0.006, z test). The autosomal dominant NDC gene set had the largest per-gene effect, with the average per-gene effect being 12.3 times the exome-wide average, though not significantly different from it due to the large standard error of the estimate. In contrast, the autosomal recessive gene set did not show an association with IQ significantly different from the exome-wide average or even from zero. The next strongest association was that of the EA GWAS gene set, with an average per-gene effect 6.6 times greater than the exome-wide average ($p=1.5x10^{-7}$, z-test), followed by the gene sets defined based on GWASs for intelligence and schizophrenia (4.6 and 3.4 times the exome-wide average; $p=2.8x10^{-4}$ and 0.048 respectively).

The differences in average rare variant effect sizes per gene between gene sets observed in could simply be driven by their different distributions of underlying selection coefficients (Extended Data

Figure 5C), since the majority of the $RVB_{pLoF}$ signal is due to variants in highly constrained genes (Extended Data Figure 6A). In order to assess the enrichment of per-gene effects in a gene set relative to similarly constrained genes, for each gene set, we simulated 100 gene sets of equal size with matching underlying selection coefficient distributions and pooled their effect estimates (see Supplementary Methods). Enrichment was defined as the ratio of the effect estimate for the original gene set to the pooled estimate from the randomly sampled gene sets. The autosomal dominant NDC gene set did not show any enrichment, suggesting that the relatively high number of evolutionarily constrained genes in this gene set is the main driver of the large per-gene effect (Extended Data Figure 5D). In contrast, the only Bonferroni-significant enrichment was observed for the EA GWAS gene set (ratio=2.47; $p=1.6x10^{-4}$), with nominal enrichments for the intelligence and prenatal gene sets and a relative depletion of signal in the non-brain gene set. We observed similar results using the composite cognitive performance measure in MCS, albeit with attenuated levels of enrichment compared to ALSPAC that were nominally significant only for the EA GWAS gene set (Figure S6). These results suggest that genes prioritized via the EA GWAS contribute more variance to the RVB associations with childhood IQ than expected given their level of constraint, and potentially also for those prioritized via the intelligence GWAS and genes preferentially expressed in prenatal brain.

Given the large enrichment and per-gene effects observed for the $RVB_{pLoF}$ in the EA gene sets, we then sought to compare how different gene inclusion criteria impact their associations on IQ in ALSPAC. We previously used a 5% FDR cutoff from the prioritized genes in Lee et al. regardless of whether the gene was the closest gene to the significant SNPs informing the prioritization. We additionally considered all combinations of using a 1% and 0.1% FDR cutoff as well as restricting to only genes that were the closest genes to the GWAS significant SNPs, resulting in six gene sets. Both when considering all genes or only closest genes, using more stringent FDR thresholds led to stronger per-gene standardized effects (Extended Data Figure 5E). When restricting to only closest genes, the point estimates of the effects increased substantially for all FDR thresholds. For example, when comparing the gene sets with an FDR cutoff of 0.1%, the per-gene standardized effects increased by 51% when restricting to the closest genes, i.e. the per-gene variance explained was 2.3 times greater, though the standard errors increased substantially due to the decrease in the gene set size. As there was modest overlap with the AD NDC gene set, we further considered the EA gene sets after excluding genes overlapping the AD NDC gene set to ensure they were not driving the associations. After doing so, the per-gene effect point estimates all became stronger though not significantly so (Extended Data Figure 5E), likely due to the relative rarity of damaging variants in the highly constrained NDC genes. These results were recapitulated using a composite measure of cognitive performance in MCS (Figure S6C). We conclude that genes prioritized via common variant GWAS of cognitive traits in adult populations converge with those that have outsized effects on IQ in early life.

#### Supplementary Note 4: Relative contributions of deleterious pLoF *de novo* and inherited variants to IQ

We sought to compare the relative contribution of inherited versus *de novo* pLoF variants in evolutionarily constrained genes (n = 3160 autosomal genes) to IQ in ALSPAC and our composite cognitive performance measure in MCS. To simplify interpretability of the influence of *de novo* variants in such genes on these phenotypes, we first compared the effect of the $RVB_{pLoF}$ previously used to the effect of the the number of evolutionarily constrained genes in which an individual carries a pLoF variant (constrained pLoF count). The standardized effect of $RVB_{pLoF}$ and constrained pLoF count on IQ were not significantly different from each other at any age in ALSPAC, though the latter consistently produced slightly weaker effects (Extended Data Figure 6A). Thus, for this analysis, we considered the associations of constrained pLoF count with measures of cognition.

We then compared the unstandardized effects of the constrained pLoF count of putatively inherited versus robust *de novo* variants on IQ in the fully exome-sequenced trios from ALSPAC (n=958) using mixed-effect modeling. The *de novo* constrained pLoF count had a larger point estimate, but it was not significantly different from that for inherited variants (Extended Data Figure 6B). However, power may have limited the ability to estimate a difference in the effects, as only 1.8% of children had at least one *de novo* pLoF in a constrained gene, compared to 12% with at least one inherited pLoF in a constrained gene. The age interaction with *de novo* constrained pLoF count was nonsignificant, though in a concordant direction with the inherited pLoF count and the total pLoF constrained count (Extended Data Figure 6B). The ratio of the main effect to the age interaction effect was highly concordant between *de novo* mutations and inherited variants (18.6 and 17.9, respectively), potentially suggesting the age attenuation of the influence of rare variants acted similarly for *de novo* and putatively inherited variants, though the error bars are large.

The variance of the constrained pLoF count for *de novo* mutations was significantly lower than that for inherited variants (0.018 versus 0.14, $p<10^{-10}$ F test). The variance in IQ explained by the constrained pLoF count of inherited variants was nearly equal to that of all variants and about four times that of the *de novo* pLoF count (Extended Data Figure 6C). As the inherited and *de novo* constrained pLoF counts were uncorrelated (r=-0.04, p=0.13) and the estimated age interaction effects were proportionally consistent, these results suggested that the inherited constrained pLoF count explained between 3.5-4 times more variance than the *de novo* constrained pLoF count in IQ across development in this cohort. Virtually identical results were observed when conducting the same analysis on cognitive performance in MCS (Figure S1).

However, this analysis has several limitations. First, our *de novo* variant calling was calibrated to maximise specificity, possibly at the expense of sensitivity, so some of the putatively inherited variants may in fact be *de novo*. Second, the fact that children in full trios in ALSPAC and MCS are biased towards coming from households with higher educational attainment (e.g. effect of maternal and paternal EA on being in a full trio in ALSPAC, odds ratio = 1.08 and 1.12 respectively, $p<10^{-15}$ for both) and older parents (maternal age at birth OR = 1.029, p=0.00653 in

ALSPAC; OR=1.078, $p=5x10^{-15}$ in MCS) likely means we have overestimated the fraction of the rare variant effect that is due to *de novo* mutations compared to an unbiased sample. This implies that the trio parents are likely to have fewer inherited rare variants reducing cognitive ability and that they probably pass on more *de novo* mutations due to having, on average, higher parental age[18,19].

#### Supplementary Note 5: Quantile regression results using cognitive ability measures in MCS and UK Biobank

As additional replication for the quantile regression results (Figure 3), we then considered the association between $PGI_{EA}$ and a single measure of cognitive performance from each of MCS and UK Biobank. In MCS, cognitive tests were administered at multiple ages, but previous work showed that these are not longitudinally invariant[47], so we instead extracted a single composite cognitive measure from the tests administered at ages 3 and 7 (see Methods) to represent overall cognitive performance in early childhood. In UK Biobank, we used the results for the verbal-numerical reasoning test (sometimes called “fluid intelligence”) conducted at baseline, to represent adult cognitive performance. We hypothesized that we would see relatively uniform effects across quantiles of early childhood cognitive performance measured in MCS, as we did for IQ in ALSPAC at age 4 (Figure S2). Given the differential age interactions across quantiles observed in ALSPAC (namely the increasing effect with age that is seen only in the top half of the distribution), we predicted that, in UK Biobank adults, the $PGI_{EA}$ effects would be markedly stronger at the top 5% and median than the bottom 5%. Our results were concordant with these two predictions (Extended Data Figure 7B): neither the population (n=5,920) nor direct (n=5,309) effects were statistically different across quantiles in MCS, while we found significant heterogeneity in UK Biobank when examining both the population effect (n=101,232) and direct effect (n=11,859), with the direct effect at the top 5% being 1.62 times greater than that at the bottom 5% (p=0.0084).

We then considered the association between the $RVB_{pLoF}$ and $RVB_{Missense}$ and measures of cognitive performance in MCS (n=5,666) and UK Biobank (n=101,232). Given the heterogeneity of effects on IQ observed at age 4 in ALSPAC (Figure S4), we hypothesized that in MCS, we would find stronger effects for $RVB_{pLoF}$ at the bottom 5% of our composite cognitive performance measure from early childhood. In UK Biobank, we predicted that the differences across quantiles would be minimal since we observed increasingly uniform effects across the quantiles by age 16 in ALSPAC (Figure S4). Our findings were concordant with these two predictions: the bottom 5% had an effect 1.82-times stronger than the median and 2.4-times stronger than the 95th percentile in MCS (p=0.019 and 0.011, respectively), while we found no significant differences in effects across quantiles in UK Biobank (Extended Data Figure 7B). Collectively, these results support our PGI and RVB findings using ALSPAC IQ measures.

#### Supplementary Note 6: Results of models including genetic measures, parental education, and perinatal exposures

We first considered the effects of genetic scores, parental education and perinatal factors (maternal health and weeks born preterm) on mean IQ in ALSPAC using mixed-effects linear regression. Since perinatal factors considered are both associated with lower parental EA[50–52], and parental EA is correlated with both the parents' and the child's genetics, the effects of these various factors on the child's cognitive ability were likely not independent. Thus, we considered a model in which we jointly fit parental EA and the perinatal exposures together with the genetic scores that showed significant effects in Figure 1 and 2, namely offspring and parental $PGI_{EA}$ and the child's $RVB_{pLoF}$ and $RVB_{Missense}$ (conditional estimates shown in orange in Figure 4A). For replication, we ran a linear regression of our composite cognitive performance measure in MCS against the genetic scores, parental education and weeks born preterm, omitting the 'maternal health' variable since the data were not readily available (Extended Data Figure 10).

In ALSPAC, incremental $R^2$ of the joint model (excluding weeks born preterm due to the lower sample size) relative to a baseline model with sex and genetic 10 PCs was 22.8% at age 4, 21.2% at age 8 (p=0.29 for a z-test for difference in variance explained compared to age 4) and 25.6% at age 16 (p=0.073 and 0.0045 relative to age 4 and 8 respectively, z test). In MCS, the incremental $R^2$ of the joint model was 16.7% relative to the baseline. We found that when jointly fit with the other variables, the parental $PGI_{EA}$ associations became nonsignificant in both cohorts, though the child's direct effect estimate did not significantly change (Figure 4A top, Extended Data Figure 10A). Similarly, the effects of the child's $RVB_{pLoF}$ and $RVB_{Missense}$ on IQ did not significantly attenuate in either cohort after controlling for these different exposure variables and $PGI_{EA}$. The association between weeks born preterm and IQ was no longer significant in the conditional analysis in ALSPAC, though this may be due to reduced power in the joint model, whereas in MCS it remained significant. Maternal and paternal EA showed similar effect sizes to each other in both cohorts, which were also similar to those captured by the direct genetic effect of the child's $PGI_{EA}$. These results suggest, apart from the parental PGIs, effect estimates were largely unaffected in a conditional analysis.

We next considered the influence of genetic scores, parental education and perinatal factors on the different quantiles of the distribution. In ALSPAC, considering each of the variables separately (Figure 4B, marginal estimates), we found that paternal and maternal education had similar magnitudes of main effects as well as uniform effects across quantiles, and we detected a nominally significant positive age interaction (p=0.029) for paternal education at the 95th percentile (Figure 4B), although the latter was not replicated in MCS, potentially due to the different demographics of this cohort. We found substantial heterogeneity in the effect of weeks born preterm in ALSPAC; this variable showed highly significant main effects at the 95th percentile (effect size = -0.120, $p=8.44\times10^{-6}$) that significantly attenuated with age (p=0.00421), as previously observed when examining the effect on mean IQ (Figure 4A bottom), while we detected no significant effects at the 5th percentile (effect size = -0.022, p=0.389). However, in MCS we did not find significant heterogeneity of effects of this variable at the tails (Extended Data

Figure 10B). This difference may be because all the ALSPAC children in this subsample were born after 32 weeks' gestation, thus excluding very and extremely premature babies who might be expected to have the greatest cognitive deficits[8]. In contrast to the prematurity result, we detected significant main effects of maternal illness at the 5th percentile ($p=5.97x10^{-8}$) in ALSPAC that attenuated with age ($p=5.06x10^{-4}$), while the main effect at the 95th percentile was nonsignificant. These results suggest that different perinatal factors may have varying impacts across the IQ distribution, with some factors predominantly affecting the upper or lower tails of cognitive ability.

We then further explored the differential effects on the different quantiles of the IQ distribution in a joint model of the genetic and other exposures (Figure 4B, conditional estimates), excluding "weeks born preterm" in ALSPAC due to high missingness. In ALSPAC, we detected significant maternal $PGI_{EA}$ main effects on the 5th and 50th percentiles of the IQ distribution (Figure 4B) which were not observed in the conditional analysis of mean IQ (Figure 4A), suggesting the heterogeneous effects across the IQ distribution may have masked these associations. This result suggests that the mother's $PGI_{EA}$, independently of its influence on the mother's actualized EA, may be associated with the child's IQ at the lower tail of the IQ distribution either due to genetic nurture or confounding tagged by the paternal $PGI_{EA}$. However, we did not replicate this finding in MCS (Extended Data Figure 10B). Maternal EA did not show heterogeneity of main effects across the IQ distribution in either cohort (Figure 4B top, Extended Data Figure 10B). However, in ALSPAC but not MCS, paternal EA did show a significant positive age interaction at the 95th percentile (Figure 4B bottom) which was not observed when considering the effect on mean IQ (Figure 4A bottom), suggesting the influence of paternal EA on IQ increases at later stages of development among those at the top of the IQ distribution. Thus, in summary, our results suggest that the relative influences of factors that best predict which children will have cognitive difficulties or will excel cognitively across childhood differ from those that best predict average IQ. However, given that some specific findings did not replicate across cohorts, there is a need to explore this in larger sample sizes.

#### Supplementary Note 7: Results of quantile regressions on genetic scores pre- versus post-imputation of IQ

The results from the quantile regressions in Figure 3 were based on IQ after imputation of missing values (Supplementary Note 1). We found that when analyzing pre-imputation IQ, some quantile regression estimates were not concordant with the observed effects post-imputation, particularly at age 4. Specifically, we found that, in cross-sectional analyses, there was no significant association between $PGI_{EA}$ and $PGI_{Cog}$ and IQ at the 5th percentile, but a large effect at the 95th percentile (Figure S2A, Table S8). However, by age 8 and 16, where the missingness was substantially lower, the pre-imputation estimates were similar to the post-imputation estimates (Figure S2A versus B). When analyzing the pre-imputation measures using mixed-effects quantile regression, this discordant effect at age 4 led to an observed positive age interaction at the 5th percentile, a nominal interaction effect at 50th percentile, and none at 95th percentile (Figure S3). However, in a direct effect analysis, the effect at the 95th percentile was greatly attenuated (Figure S3) and the age interactions generally mirrored the results observed post-imputation (Figure S3 versus Figure 3B). Specifically, when examining the direct effects, there was no age interaction effect at the 5th percentile for either PGI, but there were positive interaction effects at the 50th percentile for both PGIs and at the 95th percentile for $PGI_{Cog}$ (Figure S3). These results suggest that ascertainment bias most likely strongly impacted the PGI results pre-imputation. The fact that we observed no significant population effect of these PGIs on the 5th percentile of the distribution at age 4 may be because power has been reduced by ascertainment biases in the IQ test conducted at this age; of the subsample of participants invited to complete this test, only 81% accepted, and it may be that those families who declined were particularly biased towards lower educational attainment. Additionally, children born very prematurely were excluded from the sample and these were likely to have lower IQ. However, after conditioning on parental PGIs, the results were largely concordant with the post-imputation analysis.

The $RVB_{pLoF}$ and $RVB_{Missense}$ results pre-imputation (Figure S3-4) were qualitatively similar to the post-imputation results (Figure 3), with nominally significant larger main effects at the 5th percentile relative to the 95th and significant positive age interaction effects only detected at the lower half of the IQ distribution.
